# Quantitative multi-organ proteomics of fatal COVID-19 uncovers tissue-specific effects beyond inflammation

**DOI:** 10.1101/2022.12.21.22283785

**Authors:** Lisa Schweizer, Tina Schaller, Maximilian Zwiebel, Özge Karayel, Johannes B. Müller-Reif, Wen-Feng Zeng, Sebastian Dintner, Klaus Hirschbühl, Bruno Märkl, Rainer Claus, Matthias Mann

## Abstract

SARS-CoV-2 directly damages lung tissue via its infection and replication process and indirectly due to systemic effects of the host immune system. There are few systems-wide, untargeted studies of these effects on the different tissues of the human body and nearly all of them base their conclusions on the transcriptome. Here we developed a parallelized mass spectrometry (MS)-based proteomics workflow allowing the rapid, quantitative analysis of hundreds of virus-infected and FFPE preserved tissues. The first layer of response in all tissues was dominated by circulating inflammatory molecules. To discriminated between these systemic and true tissue-specific effects, we developed an analysis pipeline revealing that proteome alterations reflect extensive tissue damage, mostly similar to non-COVID diffuse alveolar damage. The next most affected organs were kidney and liver, while the lymph-vessel system was also strongly affected. Finally, secondary inflammatory effects of the brain correlated with receptor rearrangements and the degradation of neuronal myelin. Our results establish MS-based tissue proteomics as a promising strategy to inform organ-specific therapeutic interventions following COVID-19 infections.

## Introduction

The global SARS-CoV-2 pandemic has engendered a tremendous research effort aimed at understanding virus biology, intervening in its spread and finding therapeutic modalities. Current literature encompasses clinical observations, case studies, diverse cell culture or animal models, all of which have greatly expanded our knowledge (Huang et al., 2020; Jackson et al., 2022; Sadarangani et al., 2021). Among these efforts, omics technologies have also crucially contributed to an unbiased, systems-level view of the disease (Overmyer et al., 2021; Stephenson et al., 2021). Apart from genomic sequencing of virus variants and patients with genetic predispositions (Initiative, 2021; Lu et al., 2020; Pairo-Castineira et al., 2021), virus and host transcriptomics has added a more holistic understanding of the infection process and its consequences (Daamen et al., 2021; Kim et al., 2021a; Melms et al., 2021).

Proteomics studies gene expression at the level of functional cellular units and therefore directly measures biological functions (Aebersold and Mann, 2016). However, due to technological challenges, it has been applied much less frequently than other omics approaches. Mass spectrometry (MS)-based proteomics has enabled the investigation of the interactome between viral and host proteins, revealing potential roles of SARS-CoV-2 proteins and suggesting host factors and process for therapeutic intervention (Gordon et al., 2020; Stukalov et al., 2021). As they were performed in cell lines, these studies do not directly reflect *in vivo* conditions and do not address tissue-specific aspects of the disease. Proteomics has also successfully been employed to identify biomarkers in plasma or serum to predict patient outcomes. Several studies have analyzed a range of SARS-CoV-2 infected patients, including detailed time courses (Geyer et al., 2021; Messner et al., 2020). Main effects were related to the immune response, specifically the complement system and blood coagulation. Plasma proteomics has also complemented multiomics approaches. Interestingly, despite limited depth of quantification, MS-based proteomics had the highest predictive power in assessing disease severity (COMBAT, 2022).

By its nature, proteomics of circulating body fluids does not directly report on the effects of infection in different human organs. Tissue proteomics would be an attractive approach to investigate this, but requires post-mortem tissues and the ability to quantitatively analyze large numbers of human tissue samples. A pioneering study from the initial Wuhan outbreak analyzed seven tissues of 19 patients, which clearly highlighted the effects of virus induced inflammation and coagulation (Nie et al., 2021). These systemic effects dominated the responses in all examined tissues.

Here, we set out to develop a scalable, MS-based strategy to study the effects of SARS-CoV-2 infection on the proteomes of the major human organ systems. Taking advantage of our previous experience in the establishment of MS workflows to process formalin-fixed, paraffin-embedded (FFPE) tissue (Coscia et al., 2020; Wisniewski et al., 2013) we developed a robust and simple workflow for paraffin dissociation, sample lysis and MS data acquisition in a parallelized manner. This enabled the analysis of a post-mortem cohort encompassing more than 350 proteomes representing ten different organs. We employed proteomic quality control panels (Geyer et al., 2019), which enabled us to control for the effect of circulation-derived proteins in the measured tissue proteomes. With this approach we identified and deconvoluted the predominant, tissue-wide contribution of the systemic-inflammatory response from organ-specific effects of a SARS-CoV-2 infection. In lung tissues, COVID-19 induced damage was most extensive, with similarities and differences to other destructive lung diseases. Several tissue types showed a unique signature in response to a SARS-CoV-2 infection with the kidney and liver having most proteomic alterations apart from the lymph- and vessel system. We finally uncover secondary inflammatory effects in the immune-privileged brain involving rearrangement of neuronal receptors as well as a decrease in myelin abundance.

## Results

Our post-mortem cohort was derived from autopsies at the University Medical Center Augsburg from April to May 2020 involving 19 patients with fatal course of COVID-19 (Schaller et al., 2020). All patients had tested positive for SARS-CoV-2 by nasopharyngeal swabs and viral RNA was quantified by RT-qPCR (Hirschbühl et al., 2021). Patient age ranged from 57 to 90 years and included 74% male and 26% female patients. We also selected control tissues from a cohort of healthy donors and from patients with different types of lung disease. For each of the COVID-19 patients, we histologically assessed FFPE specimens from lung, heart, mediastinal lymph nodes, blood vessels, large vessel (aortal) walls, brain (medulla oblongata, basal ganglia), liver, spleen, kidney and adrenal glands and selected them for proteomic analysis (Figure 1A). We additionally supplemented the control group with samples from patients with different types of phenotypically similar non-COVID-19 lung diseases including influenza, non-COVID-19 diffuse alveolar damage (DAD), common interstitial pneumonia (UIP) with progressive fibrosis of the lung, and fibrosing organizing pneumonia (OFP) (Figure 1B). Altogether, this resulted in more than 350 human tissue proteomes to be analyzed in a robust, quantitative and reproducible manner. Patient characteristics including BMI, smoking status and comorbidities are listed in Supplementary Tables 1 and 2.

**Figure 1.**
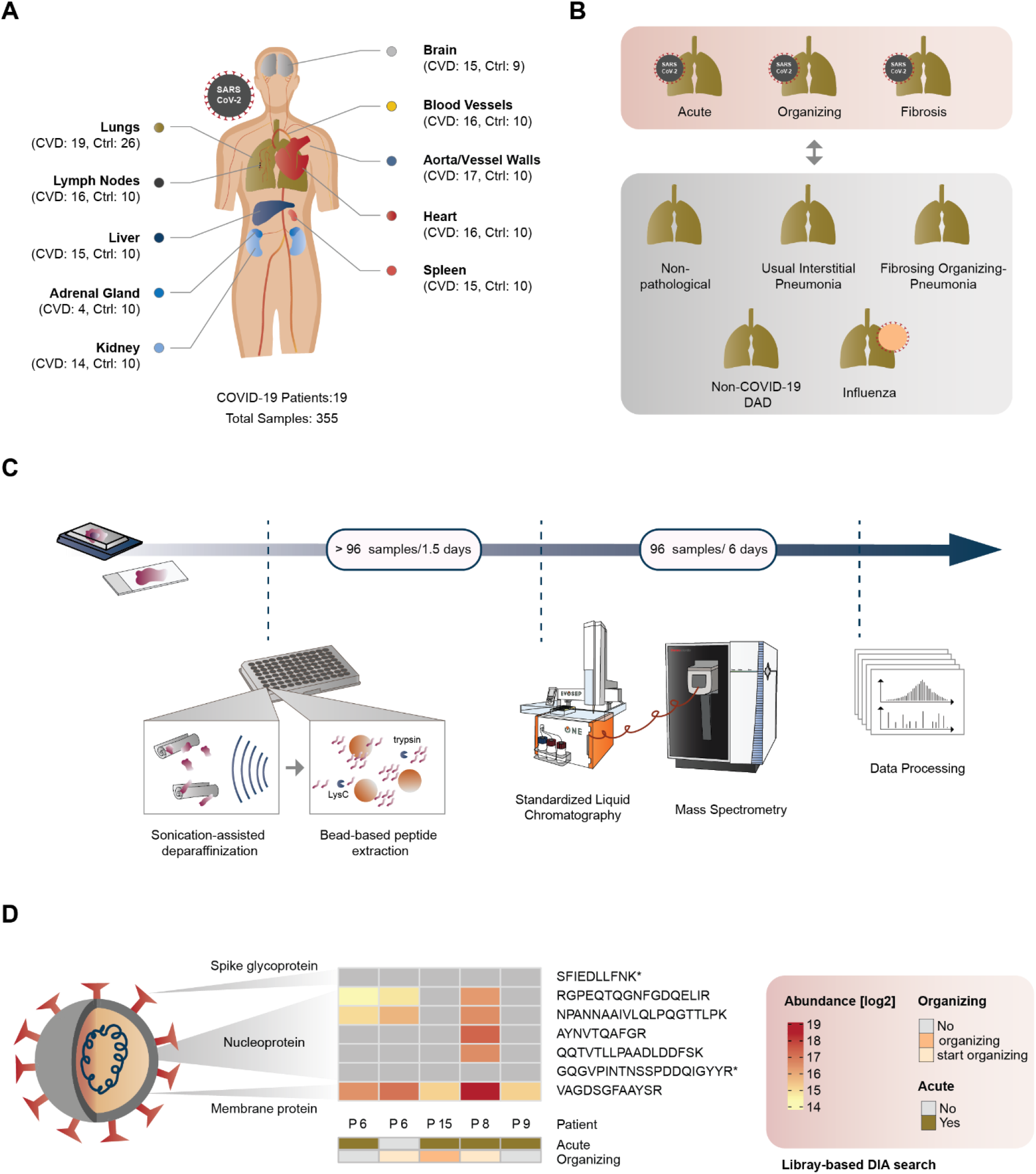
Study design and MS-based proteomics workflow overview. **A** Overview of the cohort and organs included in this study. A total of 352 specimen from 19 patients with fatal COVID-19 (CVD) were complemented with heterogeneous control groups (Ctrl) of non-pathological tissue (all tissue types) and non-COVID-19 disease phenotypes (lung tissue). **B** Selected tissue regions of the lungs were separated into acute, organizing, and fibrotic stages of COVID-19. Next to non-pathological controls, four non-COVID-19 pathologies of the lungs were compared to the different stages of the disease. **C** Schematic representation of the high-throughput and standardized proteomic workflow developed in this study. Starting from paraffinized tissue, samples were prepared by the integration of focused sonication and bead-based aggregation in a 96-well format, and analyzed by LC-MS/MS using standardized gradients in single-runs per proteome. **D** Identification of peptides from diverse components of SARS-CoV-2 in the lungs confirmed by a library-based approach. Relative abundances of peptides in each patient sample denote the presence (heat map color scale) or absence (grey) of signal in the respective sample. Annotated peptides by asterisks (*) were identified in the proteomic libraries only. The identification of these peptides in directDIA mode is shown in Supplementary Figure 1E.

### Development of a streamlined tissue proteomics workflow

Recent efforts in the acquisition of proteomic data in a high-throughput format have aimed at simplifying and streamlining the chromatographic setups (Bache et al., 2018; Bian et al., 2020; Gao et al., 2022; Messner et al., 2021). Facing the need for a rapid and universal strategy, we wanted to build on these concepts as well as previous work that allows the preparation of pathological tissue samples for direct MS analysis (Coscia et al., 2020; Foll et al., 2018; Jiang et al., 2007; Muller et al., 2020; Wisniewski et al., 2013). Briefly, to enable parallelized, non-toxic paraffin dissociation and sample lysis, we integrated protein aggregation capture (Batth et al., 2019; Hughes et al., 2019) with sonification-based and heat-assisted deparaffinization of archived FFPE tissue. Our new protocol allows all steps to occur in one well of a 96-well plate, does not use toxic chemicals and would be also be suitable for non-paraffinized tissue and complete automation. Upfront of MS analysis, we used a standardized liquid chromatography (LC) system to reduce measurement complexity while keeping with high reproducibility (Bache et al., 2018), using data independent acquisition (DIA) on an orbitrap analyzer. In combination, we found this approach to provide an appropriate methodology for a large-scale translational project based on pathologically preserved FFPE tissue proteomes while reducing the time of active tissue handling, improving reproducibility and promoting robustness (Figure 1C, Methods section).

We previously measured tissue proteomes with 2h gradients, whereas the Evosep One system features either 15 or 30 samples per day (SPD), corresponding to 44-or 88-minute gradients. For the purpose of our study, we chose the 15 SPD method as the best compromise of depth of coverage and throughput. Compared to our previous, slower LC setup, we achieved on average 13% less protein identification at 35% less measurement time (Supplementary Figure 1A,B). To also evaluate the sample processing quality of streamlined workflow, we compared it to our previous state-of-the-art protocol for the processing of deparaffinized FFPE tissue in a 96-well format (Coscia et al., 2020) by processing a subset of liver tissues from our cohort both on the same LC and MS instruments. Measured by the identification of protein groups, our greatly simplified protocol achieved equal performance (Supplementary Figure 1C).

Finally, we applied our new workflow to acquire a total of 352 tissue proteomes in less than one month of total measurement time. By using this integrative approach, we overall identified 7,315 proteins in all tissue types and demonstrated robust and repeatable identification levels in the COVID-19 and the control groups, ranging from about 2,000 different proteins in blood vessels to nearly 5,000 in lymph nodes (Supplementary Figure 1D).

### Detection of SARS-Cov2 peptides in vivo in human lung tissue

MS-based proteomics is unbiased in the sense of not being targeted to specific molecules of interest. This should make it possible, in principle, to directly detect and quantify SARS-CoV-2 virus proteins, although not reported by a previous study despite extensive fractionation of the samples (Nie et al., 2021). After adding virus protein sequences to the search database, we indeed unequivocally identified peptides of the most highly expressed viral proteins. We manually verified our findings by matching our experimental data to predictions of fragment intensities of these peptides and the extracted ion chromatograms at the MS2 level as described recently (Zeng et al., 2022) (Supplementary Figure 1E). The viral proteins were found in several lung tissue samples of four of the subjects, all of whom had either acute or organizing diffuse alveolar damage (DAD) and were among those with highest viral load as judged by qPCR (Figure 1D). These SARS-CoV-2 peptides were either derived from the membrane (M, UniProt: P0DTC5) or the nucleoprotein (N, UniProt: P0DTC9). Hence, the identification of these viral peptides provided a proof of principle of the depth and accuracy of our FFPE based workflow from post-mortem tissue. We conclude that the robustness, reproducibility and sensitivity of our workflow is of adequate quality for the purposes of our study.

### The systemic inflammatory response of COVID-19

The clinical presentation of COVID-19 is characterized by fever, worsening of the general condition and acute organ injury highlighting the systemic nature of the disease as well as the involvement of multiple organs. In order to describe these effects at the level of the proteome, we compared the differential protein expression between COVID-19 patients and respective control tissues across all organs in our study. The proteomic alterations were most substantial in the lungs, in which 25.6% of all proteins significantly changed (minimum fold-change 1.5 at a q value of 0.05, Supplementary Figure 2A). The lymphatic and blood vessel systems also had prevalent, although fewer changes (17% and 24% of proteins, respectively).

To characterize the proteome of our tissues with regard to previous reports and known properties of COVID-19, we performed a GSEA enrichment using the Reactome, KEGG and GO Biological Processes databases (Gillespie et al., 2022; Kanehisa et al., 2022; Mi et al., 2019) and extracted enriched pathways that showed substantial co-occurrence across the organs of our study (>= 6 organs, Figure 2A). Notably, these biological processes highlighted the typical systemic-inflammatory effects of the disease. The most prominent was the complement cascade, of which we quantified 13 different proteins that showed dysregulation across seven of ten different tissue types (Figure 2B). Proteins initiating the cascade, such as Complement C1q subcomponent subunits A, B and C (C1QA, C1QB and C1QC) were mainly enriched in lungs and lymph nodes. Complement factors C3, C5, C7, C9, in contrast, were commonly upregulated in lymph nodes and aorta/vessel walls.

**Figure 2.**
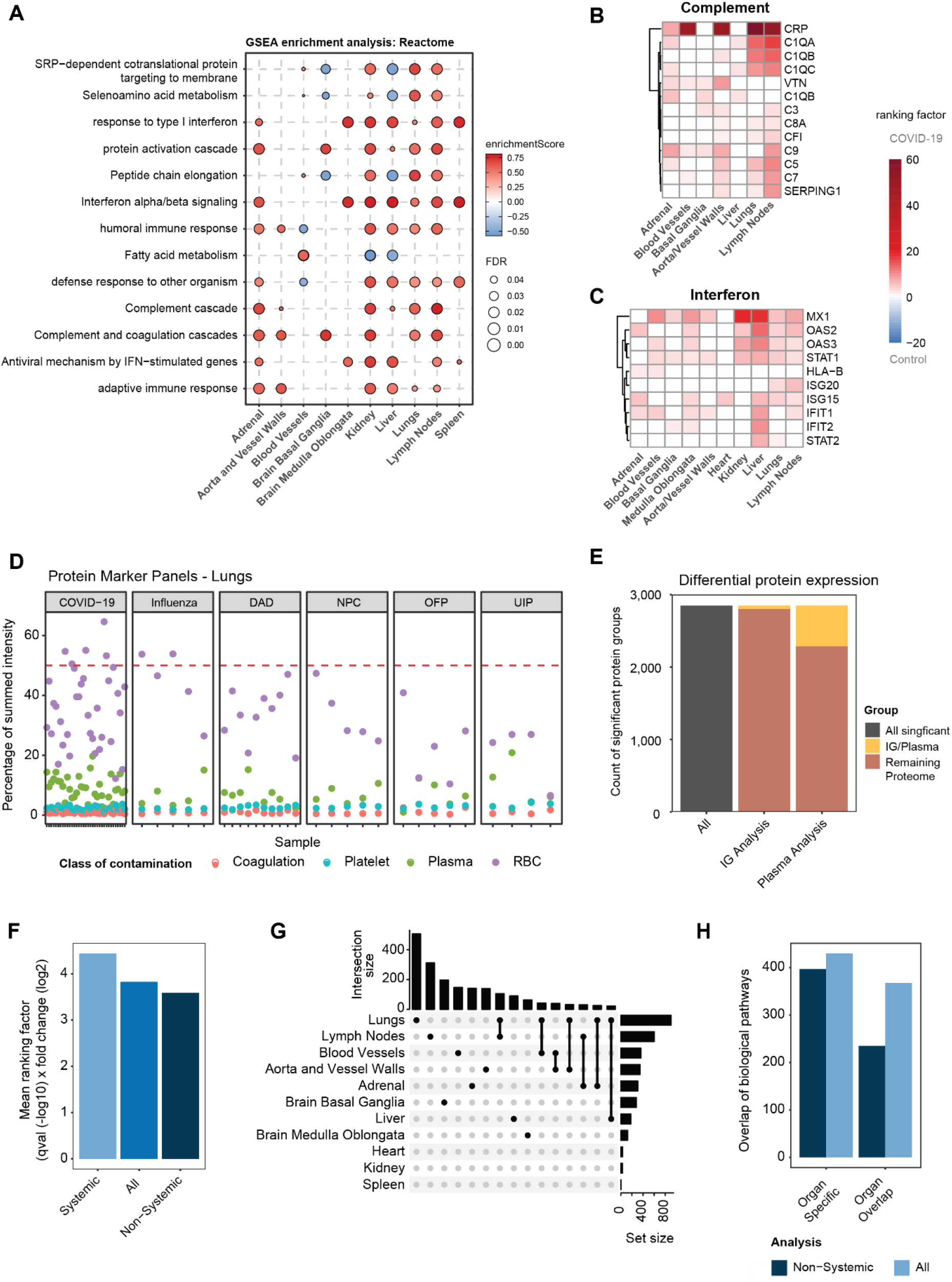
The systemic-inflammatory response masks tissue-specific effects. **A** Terms of biological pathway enrichment analysis (GSEA) of the differential protein expression between COVID-19 and controls and a minimal overlap of six organs in this study using the pathway Reactome database. Color codes of each enrichment term depict the enrichment score, whereas point size reflects the respective FDR. **B, C** Representative protein panels for the biological pathway ‘complement system’ (**B**) the ‘interferon signaling’ (**C**) with significance (fc > 1.5, q-val < 0.05) in >= 3 organs and 2 organs, respectively. Each heat map depicts the ranking factor (q-value (-log10) x fold change (log2)) for a protein in different organs as a measure of comparable significance. **D** Assessment of the prevalence for proteins in previously identified quality marker panels on coagulation, plasma, platelets and red blood cells (RBC), exemplarily shown for lung tissue. The percentage of summed protein intensities for each panel is illustrated for each sample across all disease pathologies included into our study. The ratio of 50% signal of the total protein abundance is highlighted (dashed red line). **E** Ratio of plasma or immunoglobulin-related proteins on the total of significantly differentially regulated proteins (fc > 1.5, q-val < 0.05) between COVID-19 and control samples across all organs. **F** Calculation of the mean ranking factor (q-value (-log10) x fold change (log2)) for significantly dysregulated (COVID-19 vs control) proteins of the original proteome (All) as well as proteins associated to the systemic and non-systemic effects in tissue across all organs. **G** Upset plot depicting the intersection of significantly dysregulated (COVID-19 vs control) proteins across all organs. **H** Comparison of a biological pathway enrichment for significantly dysregulated (COVID-19 vs control) proteins of the original proteome and after identification of the systemic effects using the Reactome, KEGG and GO biological process databases across all organs. For each enrichment, the number of pathways is shown for organ specificity and overlap in at least two organs.

The second most altered group consisted of components of the interferon pathway that were consistently upregulated in all measured tissue types, including brain, liver, kidney and adrenal glands (Figure 2C). This included proteins like MX1, OAS2, OAS3, and IFIT1 mediating interferon-induced and RNA-specific antiviral activity. Proteins involved in the transmission of type I interferon signaling (STAT1) or the regulation of the IFN-I response (ISG15) were also significantly higher abundant in several organs.

We noticed that the abundance of acute-phase proteins CRP and LBP as well as the predictive markers SERPINA3 and SAA1 that had been associated with severe COVID-19 disease progression in blood and plasma (COMBAT, 2022; Geyer et al., 2021; Messner et al., 2020), were also clearly increased in the large majority of organs (Supplementary Figure 2B). This was also true for the lung diseases used for comparison (Supplementary Figure 2C). CRP was upregulated in all disease phenotypes of the lungs compared to control tissue. LBP and SERPINA3 were mainly elevated in lung damage induced by influenza and non-COVID-19 DAD. Hence, host systemic inflammation upon SARS-CoV-2 infection appeared to be predominant and organ-spanning in our proteomic tissue screening, recapitulating clinical features previously reported in proteomic tissue studies of COVID-19 (Nie et al., 2021).

### Deconvolution of organ-specific effects in COVID-19

Prompted by the ubiquitous predominance of systemic effects in COVID-19, we next investigated the possible contribution of the circulation-derived proteome in our tissue-based study and its impact on data interpretation. For this purpose, we employed proteomic ‘quality control panels’ which we had previously developed to control for the effect of blood compartment contamination in the proteomes of body fluids (Geyer et al., 2016; Karayel et al., 2022). We focused on representative markers of panels for coagulation, platelets, plasma and red blood cells (RBC). This revealed that markers of the RBC panel dominated some specimen of the COVID-19 cohort and accounted for more than 50% of the summed intensity of the MS signal (exemplarily shown for lung tissue in Figure 2D).

The above results suggested that blood was present in our proteomic samples and contributed significantly to the proteomic profile of the organ samples as expected since residual blood in the preparation cannot be excluded during postmortem processing of human tissue. At the same time, circulation mediated effects of the immune system are part of the systemic effects of COVID-19 infection, including tissue destruction. To distinguish these contributions of the blood proteome, we next considered proteins that are constituent and commonly identified parts of plasma proteomes (Geyer et al., 2016) including immunoglobulins (in total 1060 different proteins, Supplementary Table 3). We termed this the ‘circulation-mediated’ proteome subset.

Mapping this subset to the significant proteomic alteration between COVID-19 and control samples across all organs, revealed that it accounted for 24.1% of all significantly differentially regulated proteins, 19.9% of which were various immunoglobulins (Figure 2E). To interpret these differences, we considered the ‘ranking factor’, defined as the product of fold-change and statistical consistency (corrected q-value), for each protein (Xiao et al., 2014). The mean ranking factor of the circulation-mediated subset was substantially higher than the mean of all significantly altered proteins (Figure 2F). The same value of the non-circulation-mediated subset was clearly decreased, indicating that overall results were indeed influenced by the blood proteome. Based on these findings, we hypothesized that the circulation-mediated proteome subset might have masked organ-specific effects in our data interpretation. Interestingly, when separating these effects, the remaining effects of COVID-19 on the host proteome became more organ specific, with the majority of tissue now exhibiting unique and distinct groups of differentially regulated proteins. As before, the number of differentially regulated proteins with unique changes was most substantial in the lungs, followed the lymph nodes and the basal ganglia of the brain (Figure 2G, Supplementary Figure 2D).

To interpret the impact of the circulation-mediated effects, we extended the analysis using GSEA pathway enrichment across all organs. Interestingly, the number of enriched pathways decreased markedly in each organ upon data deconvolution (Supplementary Figure 2E). The organ-specificity of the pathway enrichment remained nearly unchanged. However, we observed a drastic decrease of the organ overlap when considering the non-circulation-mediated proteome subset (Figure 2H).

Thus, our analysis uncovered a predominant role of the circulation-mediated effects in the COVID-19 tissue proteome data. In this regard, we recapitulate the central role of circulation-derived proteins in COVID-19 that are related to symptomatic characteristics of the disease such as destructive injury in lungs and other systemic effects during its progression. The quality marker panels that we had previously used to exclude contaminated body fluid samples here proved useful to distinguish organ-specific from systemic effects. This informed all subsequent analysis to identify the truly organ-specific damage upon a SARS-CoV-2 infection.

### Pulmonary manifestations of COVID-19

To separate non-systemic from the systemic effects in the tissue proteome changes, we implemented the deconvolution of the systemic effects in COVID-19 as described above, first focusing on the manifestation of COVID-19 in the lungs. In comparison to healthy lung tissue (non-pathologic controls, NPC), 384 and 533 proteins were up- and down-regulated in COVID-19, respectively (Figure 3A). Fibroblast growth factor receptor substrate 3 (FRS3) and the negative regulator of collagen production Reticulocalbin-3 (RCN3) were among the most up-regulated (5.4 and 3.9-fold, respectively). Interestingly, proteomics directly revealed details on the immune defense against viral infections, including drastic increase in antigen CD177, Integrin alpha-M (ITGAM) as well as the complex-forming E3 ubiquitin-protein ligase DTX3L and PARP9. In contrast, proteins providing tissue structure such as the basal cell adhesion molecule (BCAM) and the tight junction organizing protein EPB41L5 were significantly downregulated. Further downregulated proteins of functional or structural significance in the lungs were the alveolar type I (AT1) cell lineage markers AGER/RAGE, CLIC5 as well as Caveolin-1 (CAV1) and the Pulmonary surfactant-associated protein C (SFTPC) which is exclusively expressed in alveolar type II (AT2) cells.

**Figure 3.**
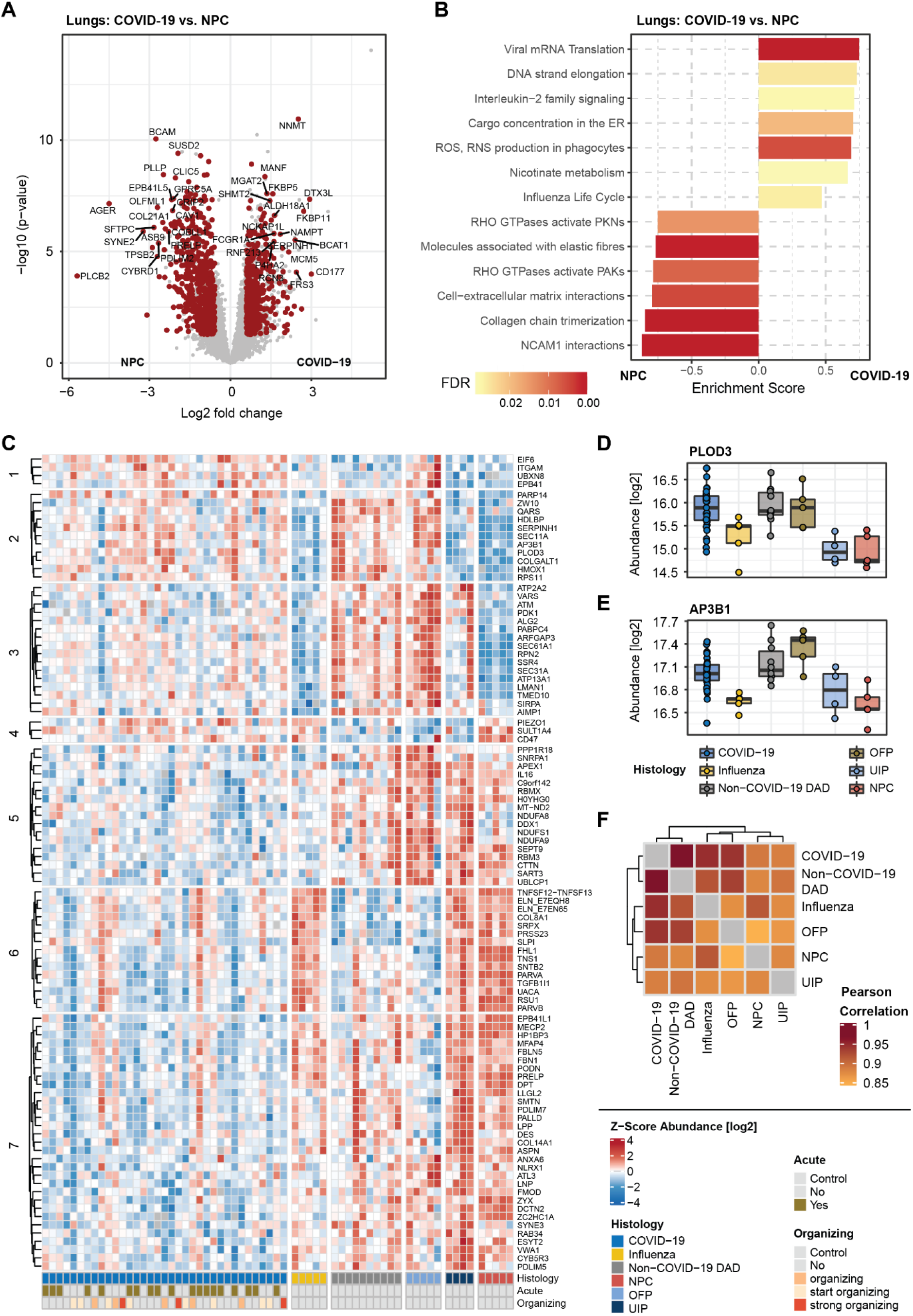
Manifestation in the lungs and classification among related lung diseases. **A** Differential protein expression between COVID-19 and non-pathological control samples (NPC). Significant proteins (t-test, q-val < 0.05, fold change >1.5) are highlighted in red. **B** GSEA enrichment for the differentially regulated proteins in COVID-19 and NPCs in **A** using the pathway Reactome database. For each protein subset the most representative terms are shown. **C** Heatmap of ANOVA significant (ct < 0.001) proteins that have been filtered for differential regulation by post-Tukey significance (adj. p-value < 0.05) in at least two lung diseases compared to COVID-19. Annotations show the groups of lung diseases as well as the disease pathology by acute or organizing tissue architecture (bottom). **D, E** Exemplary depiction of the differentiation of COVID-19, non-COVID-19 DAD and in cluster 2 (**C**) by protein the abundances of PLOD3 (**D**) and AP3B1 (**E**). **F** Pearson correlation of median intensity values across the groups of our lung cohort, indicating the overall level of similarity between COVID-19 to the other lung diseases.

We next extended the protein level analysis with a biological pathway enrichment in comparison to NPC specimen. This highlighted specific processes that divided in either enriched or downregulated processes in COVID-19 (Figure 3B). Some reflected protein assemblies directly involved in defense against the viral infection, such as phagocytic degradation and interferon signaling, as well as viral mRNA translation and protein cargo concentration in the ER induced by protein translation. Notably, we also observed increased levels of proteins related to ‘nicotinate metabolism’. Next to proteins involved in the synthesis and processing of Nicotinamide adenine dinucleotide (NAD), the nicotinamide N-methyltransferase (NNMT) was among the most significantly upregulated proteins in COVID-19 compared to control samples of the lungs. In contrast, processes associated with the maintenance of tissue structure such as elastic fiber formation and the organization of the interaction of cells with the extracellular matrix appeared substantially less abundant in COVID-19 tissue specimen, supporting the protein level results described just above. Interestingly, also the activity of RHO GTPases showed decreased effects, pointing out alterations in the cytoskeletal organization and actin/tubulin dynamics in the cell. A detailed protein composition of the respective pathway term is given in Supplementary Table 4.

### Classification of COVID-19 among lung disease of related pathology

To contrast the pulmonary manifestations of COVID-19 to other lung diseases of related pathology in addition to the NPCs, we next compared our COVID results to those from samples of patients with influenza, non-COVID-19 DAD, common interstitial pneumonia (UIP) and fibrosing organizing pneumonia (OFP). In a principal component analysis (PCA), NPC control samples and UIP separated distinctly from COVID-19 specimen of the lungs, whereas influenza and non-COVID-19 DAD partially overlapped with COVID-19 (Supplementary Figure 3A). Comparing each lung pathology to the proteome of COVID-19 lungs, we found that the number and overlap of altered proteins was most significant in comparison to UIP followed by NPC control samples, indicating major differences of COVID-19 to usual interstitial pneumonia (minimum fold-change 1.5 at a q-value of 0.05, Supplementary Figure 3B and C-F, Supplementary Table 4).

To stratify COVID-19 from other lung disease, we next performed an ANOVA (significance cutoff ct < 0.001) followed by post-Tukey testing (adj. p-value < 0.05) to ascertain differential protein regulation in at least two lung pathologies compared to COVID-19. This identified 97 significant proteins, grouped in seven different clusters (Figure 3C). Cluster 2 specifically differentiated COVID-19, non-COVID-19 DAD and OFP from influenza and UIP. It showed increased abundances of the AP-3 complex subunit AP3B1, somatic mutations of which confer higher risk for severe COVID-19 (Luo et al., 2021) as well as the collagen-associated protein PLOD3, which is associated with common pulmonary fibrosis (Shao et al., 2020) (exemplary shown in Figure 3D, E). Likewise, levels of the ER-associated protein Vigilin (HDLBP) were elevated, which has already been reported during a SARS-CoV-2 infection (Flynn et al., 2021) and procollagen galactosyltransferase 1 (COLGALT1), correlating with increased generation of collagen in lung tissue.

In the UIP group in cluster 3, levels of ATM kinase, SEC31A1 a component of COPII complexes and the mediator of COPI/COPII-based transports TMED10 were elevated. Thus, cluster 3 is likely to display shared properties of virus-induced and pneumonia-related rearrangements via the COP vesicle system within the cell.

Cluster 6 mirrored cluster 2 as it differentiated the same patient groups, however by proteins that decreased in abundance compared to NPC controls. Among those was Elastin (ELN), a major component of the extracellular matrix, along with the respiratory tract defense Antileukoproteinase (SLPI) that has previously been associated to pseudomonas-induced fibrosis due to the degradation of neutrophil elastase (Camper et al., 2016; Mecham, 2018; Weldon et al., 2009).

Cluster 7 reflected gradual downregulation of proteins across lung diseases towards COVID-19 compared to UIP and NPC controls. This included Fibrillin-1 (FBN1), Fibulin-5 (FBLN5) and Fibromodulin (FMOD) which suggested a decline of microfibrils in the extracellular matrix. The antiviral protein NLRX1 was another interesting member of this cluster that was significantly downregulated compared to the majority of patients in the other lung diseases.

Driven by the considerable differences between the lung diseases, we tested for the level of correlation between each group of our lung cohort using median intensity values (Figure 3F). The proteomic signature of COVID-19 correlated most closely with non-COVID-19 DAD (Pearson correlation 0.97). Specimen of influenza and OFP also correlated highly with the COVID-19 phenotype (0.94), whereas usual pneumonia (UIP) correlated less (0.89). Hence, our proteomic characterization enabled the stratification of COVID-19 with respect to other lung diseases, revealing overall similarity to non-COVID-19 DAD and distinct differences to influenza-related lung damage.

### Infection-induced phosphorylation signaling in the lungs

To investigate how SARS-CoV-2 impacts intracellular signaling cascades in the lungs, we performed quantitative phosphoproteomics, using a recently described direct DIA workflow employing a FAIMS library (Stukalov et al., 2021). Here we analyzed FFPE tissue samples from 16 COVID-19 and 20 control samples including representative specimen from all previously described non-COVID-19 pathologies. Overall, we identified 11,531 phosphorylation sites on 2,681 proteins which were filtered stringently to achieve up to 2,000 sites of high confidence in each group and equal identification levels among all groups of our cohort (Supplementary Figure 4A). Compared to COVID-19, the phosphoproteomes of NPC, UIP and OFP showed significant changes (t-test, q-val < 0.05, fold change >1.5, Supplementary Table 5), which had also shown the largest differences at the proteome level (Figure 4A, Supplementary Figure 4B-D).

**Figure 4.**
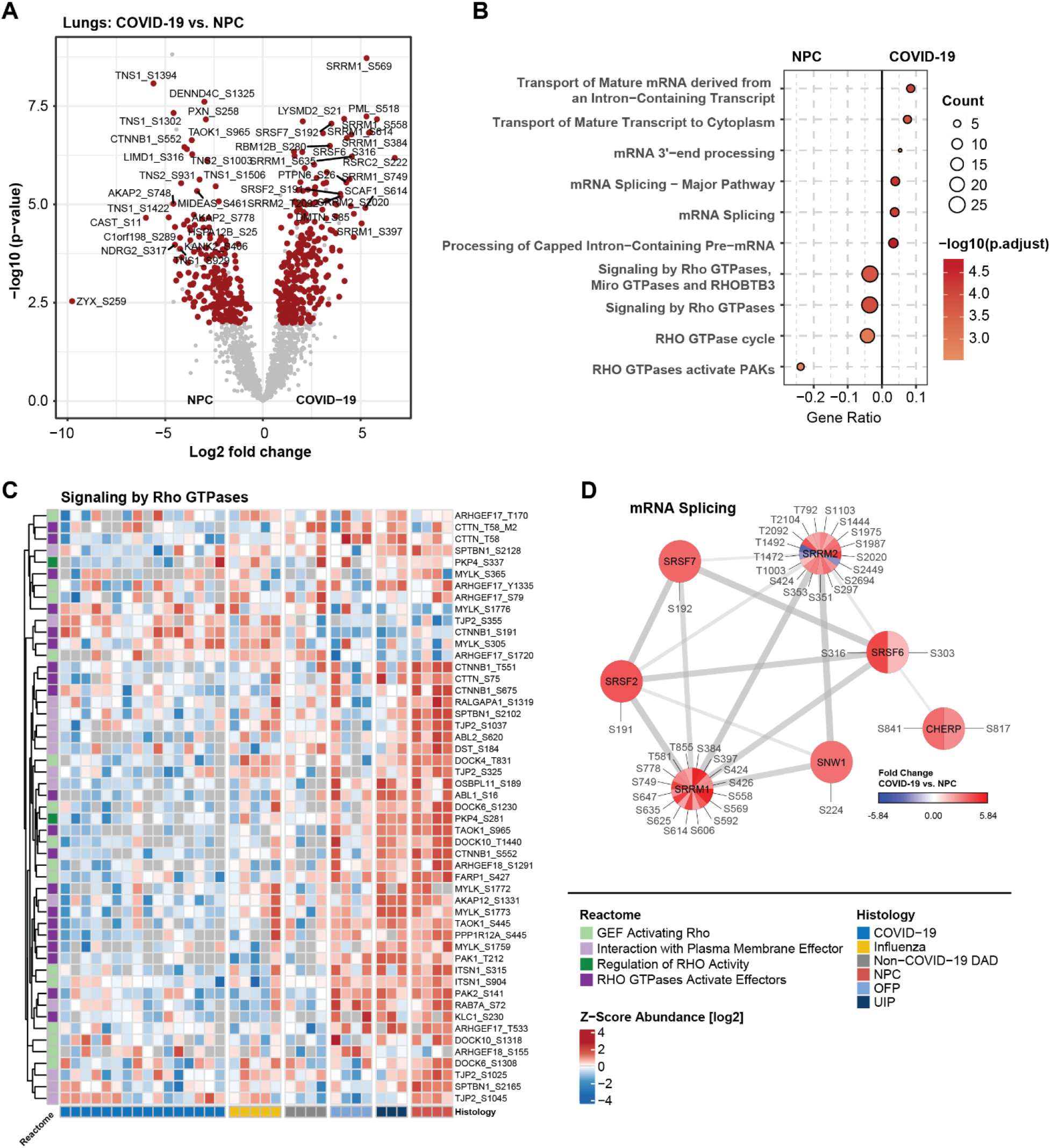
Infection-induced phosphorylation signaling in the lungs. **A** Differential phospho-site regulation between COVID-19 and non-pathological controls (NPC). Significant proteins (t-test, q-val < 0.05, fold change >1.5) are highlighted in red. **B** Biological pathway enrichment (ORA) for the differentially regulated proteins in COVID-19 and NPCs in using the pathway Reactome database. The count of proteins associated to each term is indicated by point size. **C** Heatmap of proteins involved in signaling by Rho GTPases in **B**. Annotations show the groups of lung diseases (bottom) and biological roles in the Rho Pathway (left). **D** Network representation of proteins with differentially regulated phosphorylation sites in the context of mRNA Splicing indicating their biological interaction. The color coding of each site visualizes the fold change between COVID-19 and NPC.

Reactome biological pathway enrichment analysis of significantly changed phosphorylation sites between COVID-19 and NPC revealed two main processes (Figure 4B). The first was a substantial downregulation of Rho GTPase signaling in the proteome of COVID-19 (Figure 4C), mimicking the downregulation of this pathway at the proteome level, although the phosphoregulation was additive to this (Supplementary Figure 4E,F). This involved Guanine nucleotide exchange factors (GEFs) for the regulation of Rho GTPase activity such as the dedicator of cytokinesis proteins DOCK4, DOCK6 and DOCK10 suggesting a central role of the DOCK protein family which was already shown for DOCK2 in a genome-wide association study (GWAS) in patients with severe cases of COVID-19 (Namkoong et al., 2022). Additionally, downstream phosphorylation events of Rho GTPase signaling were decreased, one average 4.3-fold. These included central kinases such as PAK1/2, the Wnt signaling pathway component Catenin beta-1 (CTNNB1) as well as the plasma membrane effectors such as the small GTPase RAB7A, the latter regulating the expression of ACE2 on the cell surface in dependence on its phosphorylation status (Daniloski et al., 2021).

The second major process affected by virus infection was related to mRNA processing and splicing. Here, we noted the hyperphosphorylation of the serine/arginine repetitive proteins SRRM1 and SRRM2 playing a major role in pre- and post-splicing (Figure 4D). SNW1, a component of the spliceosome, and the S/N-rich splicing factors SRSF2, SRSF7 and SRSF6 likewise showed increased phosphorylation. These results suggest that SARS-CoV-2 modulates host splicing machinery by acting on phosphorylation, presumably to facilitate virus replication. Notably, the increase in phosphorylation of the mRNA processing and splicing machinery was additive to the significant changes in protein levels (Supplementary Figure 4F) and were consistent with results from our recent multi-omics study of SARS-CoV-2 studying altered phosphorylation in a lung-derived human cell line (Stukalov et al., 2021).

### Organ-specific effects in COVID-19

Having analyzed the distinctive proteome alterations in the lungs, we next evaluated the organ-wide response upon SARS-CoV-2 infection beyond its entry portal. We first focused on the lymph-blood vessel system as it was the second most affected tissue (Figure 2G, Supplementary Table 6), distinguishing between lymph nodes and the walls of large blood vessels such as the aorta. Between the lymph nodes of COVID-19 and controls, 252 proteins were upregulated and 366 downregulated, after separating the organ-specific from the systemic effects as described above (Figure 5A). The DNA replication licensing factor MCM2-7 as well as the Ribonucleoside-diphosphate reductase subunit M2 (RRM2), which has an essential role in DNA synthesis, were consistently upregulated by a mean of 3.5-fold. In addition, we noted the upregulation of the nuclear importer protein KPNA2 that is targeted by the SARS-CoV-2 protein ORF6 resulting in nuclear exclusion of STAT1, thereby preventing antiviral signaling (Miyamoto et al., 2022). In contrast, proteins such as the RNA-binding protein 3 (RBM3) that suppresses the activation of innate lymphoid cells in the lungs showed major downregulation (Badrani et al., 2022).

**Figure 5.**
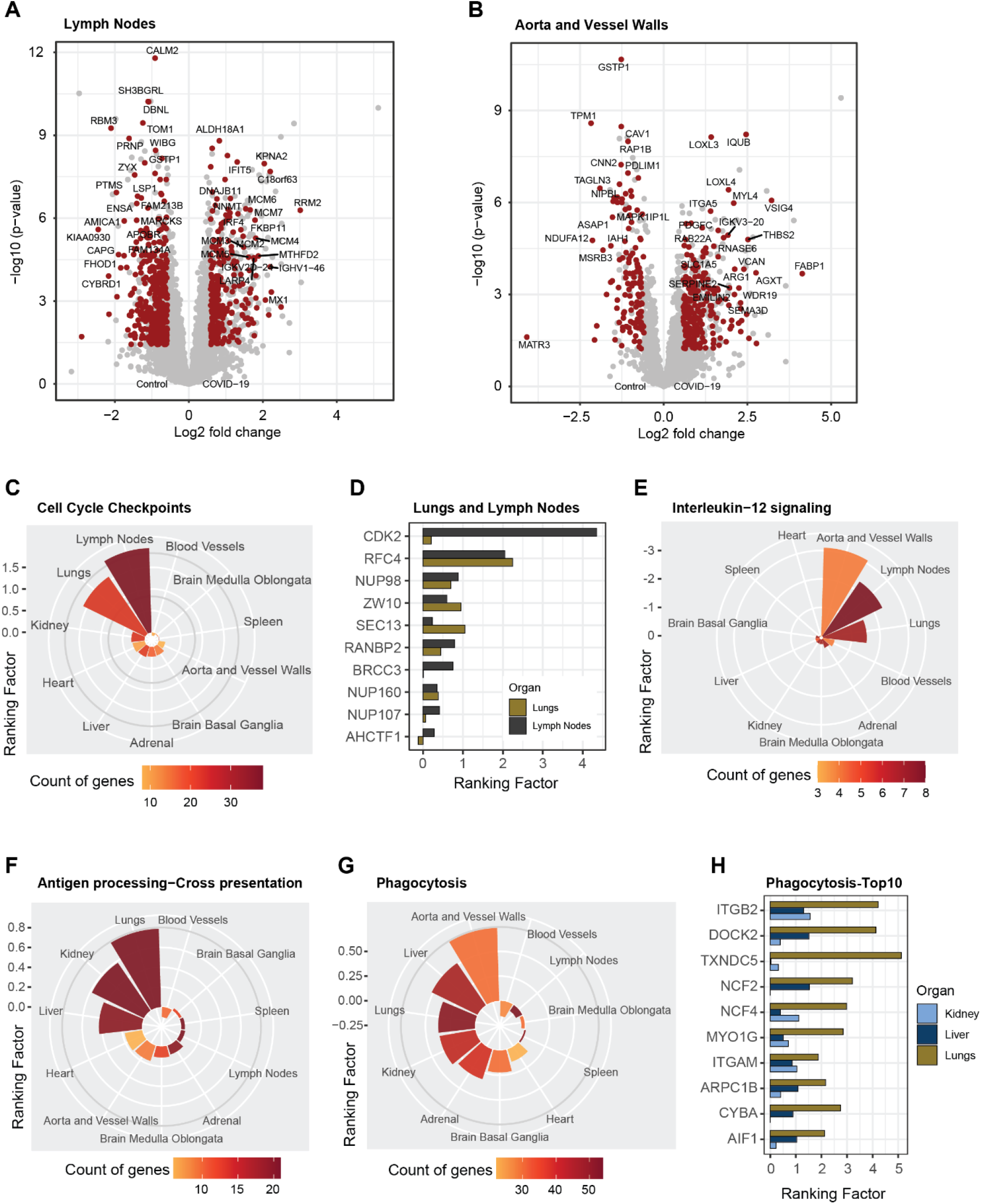
Organ-specific effects in COVID-19. **A, B** Differential protein expression between COVID-19 control specimen in the lymph nodes (**A**) and aorta/vessel walls (**B**). Protein significance (t-test, q-val < 0.05, fold change >1.5) is highlighted in red. **C** Mean ranking factor (q-value (-log10) x fold change (log2)) of all proteins associated to the Reactome term ‘Cell Cycle Checkpoints’ across all organs of this study. The number of genes identified for each organ is color coded. **D** Bar plot representing the ranking factor values for proteins from (**C**) identified in common for lungs and lymph nodes. For clarity proteasomal subunits and MCM proteins (shown in Supplementary Figure 5A) are excluded. **E-G** Mean ranking factor values depicted as in (**C**) for proteins associated to Interleukin-12 signaling (Reactome database, **E**), antigen processing-cross presentation (Reactome database, **F**) and phagocytosis (GO biological processes, **G**). **H** Bar plot representing the ranking factor values for the top 10 significant proteins from (**G**) by ranking factor identified in common for lungs, liver and kidney.

Biological pathway enrichment revealed processes such as ‘cell cycle checkpoints’ as upregulated in the lymph nodes which to less extend also occurred in the lungs (Figure 5C, Supplementary Table 6). In both organs - apart from the MCM proteins (Supplementary Figure 5A) - this was caused by proteins such as the Replication factor C subunit 4 (RFC4) and the nuclear pore complex protein NUP98, the latter being a direct target of viral ORF6 to prevent interferon signaling as shown for KPNA2 above (Miorin et al., 2020) (Figure 5D).

Cyclin-dependent kinase 2 (CDK2) was also upregulated, but this was specific to the lymph nodes rather than in the lungs (Figure 5D). RecQ-like DNA helicase BLM, the cell cycle checkpoint control protein RAD9A, the origin recognition complex subunits ORC 3/4 and the Nucleoporin Nup37 among others were exclusively enriched in the lymph nodes (Supplementary Figure 5B). Interestingly, interleukin-12 signaling was strongly decreased in lymph nodes (represented by eight effectors) as well as to less extend in the lungs and aorta/vessel walls (Figure 5E, Supplementary Figure 5C). Among the proteins of the aorta/vessel walls, the downregulation of Matrin-3 (MATR3) and Ras-related protein Rap-1b (RAP1B) related to a decreased capacity to maintain endothelial cell polarity and vascular integrity (Lakshmikanthan et al., 2014; Quintero-Rivera et al., 2015) (Figure 5B). Conversely, the regulator of inflammatory response LOXL3, the phagocytic receptor and regulator VSIG4 and platelet-derived growth factor C (PDGFC) were significantly upregulated in the proteome of COVID-19, indicating an active inflammatory response and wound healing as well as fibrotic disease.

We next investigated the effect of COVID-19 on the inner organs of the abdomen. Biological pathway enrichment revealed an increased abundance of proteins associated with the Reactome term ‘antigen processing-cross presentation’ in the kidney and liver apart from the lungs (Figure 5F). This was due to proteins such as the antigen peptides transporters 1 and 2 (TAP1/2) as well as the associated Tapasin (TAPBP) - core components of MHC class I antigen presentation. Of note, SEC61A1, a component of the translocon for the transport of polypeptides into the endoplasmic reticulum, was much less increased in the investigated organs compared to the lungs (Supplementary Figure 5D). Indicating an active response to the antigen presentation, ‘Phagocytosis’ was among the major upregulated processes in lungs, liver and kidney (Figure 5G). Proteins in this term included the integrins ITGAM and ITGB2, indicating enhanced adhesion of immune cells, as well as Cytochrome b-245 light chain (CYBA) and the neutrophil cytosol factor 4 (NCF4) promoting the phagocytic oxidative burst (Figure 5H). Pathways uniquely enriched in the liver suggested a loss-of-function effect through decrease in cholesterol biosynthesis, cytochrome P450 or glucuronidation (Supplementary Figure 5E). Interestingly, we noted a reduced expression of valacyclovir hydrolase (BPHL) in the kidney of COVID-19 patients, which is responsible for activation of anti-viral nucleoside analogs such as acyclovir (Supplementary Figure 5F).

### Secondary inflammatory effects of COVID-19 in the brain

As some symptoms of COVID-19 infection indicate neurological damage, we aimed to investigate the proteome of the brain in the post-mortem stage upon SARS-CoV-2 infection. To provide two contrasting examples of the brain by location and functionality, we analyzed specimen derived from the basal ganglia and the medulla oblongata in the brainstem (Figure 6A). In both regions we quantified more than 4,000 protein groups that showed equal levels in both areas of COVID-19 and control specimen (Figure 6B). Although we did not detect viral peptides – concordant with PCR-based analysis of the same specimens (Hirschbühl et al., 2021) – there was strong evidence of inflammation. This was specifically evident by proteins of the interferon signaling cascade such as the signal transducer and activator of transcription STAT1, the interferon-induced enzyme OAS3 as well as the ubiquitin-like protein ISG15 that showed significantly increased protein levels in COVID-19 (Supplementary Figure 6A). While the upregulation of interferon-related effectors was shared with the majority of organs, there were also specific changes in the brain.

**Figure 6.**
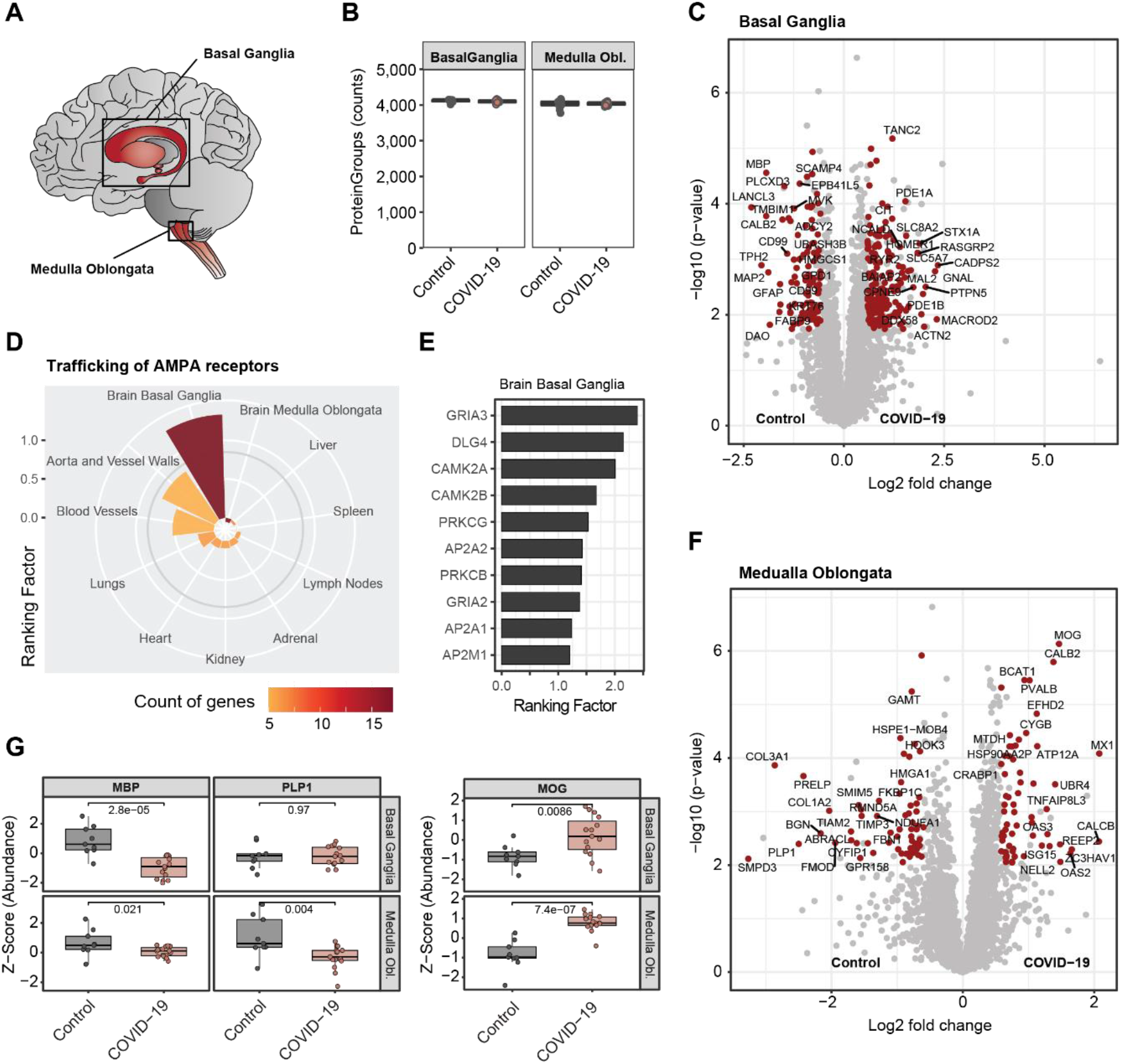
Secondary inflammatory effects of COVID-19 in the brain. **A** Brain regions selected for proteomic analysis. **B** Number of identified protein groups in the basal ganglia and medulla oblongata for COVID-19 and control specimen. **C** Differential protein expression between COVID-19 control specimen in the basal ganglia. Protein significance (t-test, q-val < 0.05, fold change >1.5) is highlighted in red. **D** Mean ranking factor (q-value (-log10) x fold change (log2)) of all proteins associated to the Pathway term ‘Trafficking of AMPA receptors’ (Reactome database) across all organs of this study. The number of genes identified for each organ in color coded **E** Bar plot representing the ranking factor values for proteins from (**D**) identified in the basal ganglia of the brain. **F** Differentially expressed proteins between COVID-19 and controls of the medulla oblongata as shown in (**C**). **G** Z-Score protein abundance of myelin-associated proteins which were derived from (**C**) and (**F**) for COVID-19 and controls in the brain. An assessment of statistical significance (unpaired t-test) is annotated for each comparison.

For the basal ganglia, these biological processes were related to an increased activity of neurotransmitter release cycles (Supplementary Figure 6B). Specifically, this included substantial upregulation of TANC2, an mTOR regulator present in neurons (Kim et al., 2021b), Syntaxin-1A (STX1A) that mediates the release of neurotransmitters from the synapse and the postsynaptic protein HOMER1 that interacts with RYR2 which was also upregulated (Figure 6C). GNAL, a signal transducer in the olfactory tract and the basal ganglia, was up-regulated. In contrast, cell surface proteins that promote and regulate the adhesion to T-cells (CD99 and the Ubiquitin-associated and SH3 domain-containing protein B (UBASH3B)) showed lower abundances. While having a minor role in the medulla oblongata, ‘Trafficking of AMPA receptor’ was specifically upregulated in basal ganglia, based on AMPA-selective glutamate receptor 3 (GRIA3) as well as both subunits of the Calcium/calmodulin-dependent protein kinase (CAMK2A/B) that regulate the signaling and trafficking of AMPA receptors by phosphorylation (Kristensen et al., 2011) (Figure 6 D,E). This suggests upregulation of AMPA receptors by inflammation (Guan et al., 2003).

GSEA enrichment in the medulla oblongata indicated increased levels of mitochondrial protein translation. Conversely, Rhodopsin-like receptors and collagen biosynthesis showed negative regulation in COVID-19, the latter indicating a loss of the stabilizing matrix in the brain (Supplementary Figure 6C). Notably, also the Zinc finger CCCH-type antiviral protein 1 (ZC3HAV1) was highly expressed in COVID-19 (Figure 6F). Reflecting the downregulation of Rho GTPase signaling in the phosphoproteomic analysis of the lungs, we found reduced levels of Biglycan (BGN) and the Rho guanine nucleotide exchange factor TIAM2.

Interestingly, the myelin proteolipid protein (PLP1) and the neutral sphingomyelinase 2 (SMPD3) were among the most down-regulated proteins in COVID-19 of the medulla oblongata (5.6-fold and 9.6-fold). This finding was complemented by the substantial downregulation of the myelin basic protein (MBP) in both regions of the brain. In contrast, myelin-oligodendrocyte glycoprotein (MOG) was among the most significantly upregulated proteins of COVID-19 in the medulla oblongata, which correlates with higher levels of MOG antibodies seen in circulation in case reports of COVID-19 (Durovic et al., 2021; Johnsson et al., 2022) (Figure 6G). As myelin protein plasmolipin (PLLP) was also downregulated in the lungs of COVID-19 (Figure 3A), this may be part of secondary inflammatory damage of myelin components in the CNS and other organs.

## Discussion

COVID-19 is a primarily a lung disease but is also associated with profound systemic inflammation in reaction to the viral infection with SARS-CoV-2. To date, numerous studies have investigated the physiological manifestation of COVID-19 and the mechanisms of SARS-CoV-2 infection. However, its effects on the different tissues in the host organism is not yet entirely understood especially at the protein level, the primary mediators of biological processes. Taking advantage of autopsies from a pioneering cohort of the early pandemic phase in 2020 at the University Medical Center Augsburg (Schaller et al., 2020), we set out to apply state of the art MS-based proteomics to investigate systemic and organ-specific effects of viral infection on the host. We obtained samples from 10 different tissues of 19 post-mortem donors, resulting in more than 350 different tissue samples. To enable timely analysis of these FFPE samples, we developed a protocol that overcomes previous limitations of throughput and sample processing by reducing workflow complexity. Using our fast, parallelized and non-toxic workflow, we identified more than 7,000 proteins in a robust and quantitative manner as a basis to understand and classify the host proteome of COVID-19. The identification of SARS-CoV-2 peptides in the lungs of severe COVID-19 patients – something not reported for unbiased proteomics in the literature before - validated the depth and specificity of our technology as well the stability of proteins in a pathology relevant matrix. No peptides were identified beyond the lungs, consistent with the limited viral spread detected by RT-qPCR in the same subjects (Hirschbühl et al., 2021).

Compared to other proteomics studies, our results confirm the global effects of inflammation seen in both plasma studies (Geyer et al., 2021; Messner et al., 2020) as well as a previous report on patient tissue (Nie et al., 2021). Although most substantial in the lungs, we observed organ-wide occurrence of systemic inflammation in COVID-19 represented by processes such as the complement cascade and interferon signaling and by central markers of inflammation such as the acute phase proteins CRP and LBP1. However, given the overlap inflammatory effects between plasma and all tissues in previous and our work, we suspected that the circulation-derived proteome masked organ-specific effects. This could be caused by blood contamination of tissues on the one hand, or the inflammatory response mediated by the circulation in the tissues. Beyond these direct and uniform effects, each tissue should also have its own specific response, mediated by tissue resident immune action (Farber, 2021). Hence, we concluded that the truly organ-specific damage upon a SARS-CoV-2 infection had to be separated from the blood proteome associated one to better understand tissue-specific effects in COVID-19. To do this, we made use of our previous work defining panels of different classes of proteins in body fluids (Geyer et al., 2016; Karayel et al., 2022). This analysis strategy indeed efficiently unmasked the tissue-specific effects of COVID-19.

The resulting data reconfirmed the lungs as most affected organs of the disease. It directly associated the characteristic fibroblastic proliferation with increased levels of the fibroblast growth factor receptor substrate FRS3 and the negative regulator of collagen production RCN3 (Martinez-Martinez et al., 2017). Conversely, decreased levels of lineage markers for alveolar type I and II cells reflected the destruction of the native cellular composition in the lungs at the protein level, also confirming previous work on the single cell RNA level (Melms et al., 2021). These markers also included AGER/RAGE, for which decreased abundances in serum of patients with severe disease progression was reported before (Yalcin Kehribar et al., 2021). This indicates a connection between stage of cellular loss in the lungs and this potential liquid biomarker. Our data further reveal that the loss of alveolar cells was accompanied by changes in the level of proteins indicating the destruction of the cellular environment and elastic fiber formation.

In comparison to other lung diseases of related pathology, COVID-19 showed highest similarity to non-COVID-19 diffuse alveolar damage (DAD), and least to usual pneumonia (UIP). COVID-19 together with non-COVID-19 DAD and OFP differed from influenza and UIP by profound collagenous fibrosis and germline variance observed for severe disease progression, such as for AP3B1 (Luo et al., 2021). The COVID-19 containing group also downregulated proteins involved in the generation of elastin polymers and microfibrillar structures suggesting a decline of the extracellular matrix. Furthermore, it showed decreased levels of the antiviral protein NLRX1 which is targeted by SARS-CoV-2 protein interaction via ORF9c (Gordon et al., 2020). Except influenza, all pathologies featured increase in cellular transport and COPI/COPII vesicles.

Analysis of peptide phosphorylation patterns in SARS-CoV-2 infected cells in culture revealed major rewiring of signaling pathways by the virus (Bouhaddou et al., 2020; Stukalov et al., 2021). Our data show that major themes of these perturbations are reflected post-mortem in FFPE tissues. This includes distinct downregulation of Rho GTPase signaling indicating central alterations in the organization of the cytoskeleton on the level of regulating kinases, such as PAK1/2, as well as downstream pathways. Furthermore, the upregulation of phosphorylation events in the context of RNA splicing support *in vitro* studies describing functional splicing as essential requirement for the life cycle of SARS-CoV-2 (Bojkova et al., 2020). In contrast, the role of SR proteins as regulators of cellular splicing during infection has only been reported in the context of HIV-1 viral replication and release so far (Wojcechowskyj et al., 2013).

Beyond the lungs, our data reflected the substantial histopathologic changes of mediastinal lymph nodes that we reported earlier (Hirschbühl et al., 2021). This study extends this finding to the molecular level, showing increased levels of SARS-CoV and SARS-CoV-2 specific antiviral signaling by proteins such as the nuclear importer KPNA2 and NUP98 in the lymph nodes (Frieman et al., 2007; Miorin et al., 2020; Miyamoto et al., 2022). Shared upregulation of the DNA replication machinery between lymph nodes and lungs, such as MCM proteins, suggests similarities in the induction of organ responses upon viral infection. In contrast to central mediators of inflammation such as interleukin-6 (Rubin et al., 2021), interleukin-12 signaling was specifically downregulated in the lymph nodes in severe COVID-19 patients (Figure 5E) supporting reports of higher interleukin-12 abundance in serum of mild compared to severe disease (Tjan et al., 2021). Beyond the lymph nodes, endothelialitis has been reported early in the pandemic (Varga et al., 2020). Our data mirror this finding by the upregulation of inflammatory and phagocytic mediators as well as corresponding indicators of vascular damage.

Inner organs such as the liver, kidney, and spleen rarely show infection with SARS-CoV-2 in histopathology or RNA sequencing. It was therefore of special interest that our proteomics results indicate active involvement of kidney and liver in the immune response of the host, mainly shown by antigen presentation and the corresponding response by phagocytosis. Decreased liver functionality has been reported in severe COVID-19 (Mao et al., 2020) and this is supported by our protein level data.

Direct invasion of SARS-CoV-2 into the central nervous system and morphological changes during disease progression in the brain are controversially discussed (Douaud et al., 2022; Hirschbühl et al., 2021; Matschke et al., 2020; Schwabenland et al., 2021). Here, we confirm highly prevalent interferon signaling in the basal ganglia and medulla oblongata on the proteomic level. In the basal ganglia, the inflammatory signature was accompanied by substantial proteomic alterations in synapses such as increase in proteins involved in the release of neurotransmitters and those associated with trafficking of AMPA receptors. Altered processes in the medulla oblongata included a loss in the stabilizing extracellular matrix and decrease of Rho signaling, the latter shared with the lungs. Most strikingly, proteins associated with the structure and maintenance of myelin were substantially reduced (Figure 6G). Thus our proteomic results directly document the consequences of secondary inflammation in the brain, correlating with the neuronal symptoms that have been extensively described COVID-19 (Venkataramani and Winkler, 2022). Degradation of myelin sheaths of the neuronal system is accompanied by inflammatory processes closely related to aging in the brain (Kaya et al., 2022; Safaiyan et al., 2016), which in turn is increasingly associated which COVID-19 (Mavrikaki et al., 2022).

In summary, our proteomics workflow allowed the in-depth profiling of 352 tissues of a post-mortem COVID-19 and control cohort. Our results recapitulated many of the findings already reported in the literature but added a plethora of quantitative molecular insights. Beyond supporting or questioning others, our resource gives rise to new hypotheses that can be validated or used in translational context by the community.

## Data Availability

All data will be publicly available upon publication in a journal.

## Acknowledgements

We thank the members of the group for Proteomics and Signal Transduction at the Max Planck Institute of Biochemistry, Martinsried, for their constant support and collaborative spirit. Specifically, we would like to thank Sophia Steigerwald, Patricia Skowronek, Ankit Sinha, Sophia Doll, Philipp Geyer, Peter Treit, Florian Rosenberger, Constantin Ammar and Thierry Nordmann for contributing valuable ideas to this study. The study was funded by the Max Planck Society for the Advancement of Science, the German Registry of COVID-19 Autopsies (www.DeRegCOVID.ukaachen.de) and the Bavarian Ministry of Science and the Arts. LS was supported by the International Max Planck Research School for Life Sciences – IMPRS-LS.

## Author contributions

LS, RC, JM-R and MM designed the study. TS, RC, KH, SD and BM collected and prepared autopsy samples. LS, JM-R, MZ and OK performed proteomic sample preparation and MS data acquisition. LS, WFZ, MZ and OK evaluated the data. RC and BM provided clinical expertise for data interpretation. LS and MM wrote the manuscript and RC provided major edits. All authors reviewed and provided feedback on the manuscript.

## Methods

### Study Design

From April 4 to May 13, 2020, 19 autopsies (15 full and 4 through limited infrasternal access) were performed. 18 patients died at the University Medical Center Augsburg (UKA), representing 86% of all Covid-19 deceased at the UKA during the first wave of the pandemic. All cases were tested positive for SARS-CoV-2 by nasopharyngeal swabs during the clinical course and postmortem. SARS-CoV-2 RNA was quantified by RT-qPCR as previously described (Hirschbühl et al., 2021). Before autopsy, informed consent from next of kin was obtained. Clinical data (including medical history, comorbidities, medication, and treatment) were obtained from electronic medical records. This study was approved by the internal review board of the UKA (BKF No. 2020-18) and the ethics committee of the University of Munich (Project number 20-426, COVID-19 registry of the UKA).

### Tissue samples

Tissue samples from organs (lungs, heart, vessels, spleen, kidneys, and brain) were immediately fixed in buffered 10% formalin solution after harvesting. After fixation (at least 12 days – 12 to 15 days), representative formalin-fixed and paraffin embedded samples (FFPE) were generated. FFPE tissue was cut (10 μm) and mounted on common microtome slides (TOMO IHC Adhesive Glass Slide, TOMO 1190, Matsunami, Japan) or membrane slides (1.0 PEN, Prod. No. 415190-9041-000, Zeiss, Germany). Lung tissue was deparaffinized and stained by Hematoxylin and Eosin for pathology assessment.

### Sample preparation

Patient material was collected in a defined area of 25 mm^2^ from tissue slides and transferred into a 96-well plate (AFA-TUBE TPX Plate, Prod. No. 520291, Covaris, Brighton, U.K.). For each plate, computer-based randomization of samples derived from patients with SARS-CoV-2 and their controls was used to exclude batch effects during sample preparation. Paraffinized and H&E-stained tissue was processed equally using Adaptive Focused Acoustics® (AFA®, Covaris) sonication and adapted Protein Aggregation Capture (PAC) (Batth et al., 2019), herein in combination termed APAC. In brief, we accomplished tissue lysis and, if present, paraffin dissociation, by sample heating and sonification. Proteins were subsequently extracted from the solution by precipitation on magnetic particles and purified by extensive washing at 50°C in isopropanol. Tryptic digestion yielded deparaffinized peptides that were finally subjected to MS-measurement.

In detail, 40 μL of tissue lysis buffer (truXTRAC Proteins - Tissue Lysis Buffer, Prod. No. 520284, Covaris) were added to each sample aliquot, followed by a pre-incubation for 10 min at 90°C in a PCR cycler. Subsequently, samples were sonified in the 96-well layout for a total duration of 300 sec per column (LE220-plus Covaris Focused-ultrasonicator, Covaris) and defined parameters (peak Power: 450.0, duty factor: 50%, cycles: 200, average power: 225). Tissue lysis was resumed during an incubation for 80 min at 90°C in the PCR cycler and concluded by repeated sonification. Enclosing, proteins were reduced and alkylated by 5 mM DTT and 20 mM CAA final concentration for 20 min at 1400 rpm and room temperature, respectively. In preparation for Protein Aggregation Capture, magnetic carboxylate modified particles (Sera-Mag™, Prod. No. 24152105050350, GE Healthcare/ Merck KGaA, Darmstadt, Germany) were washed trice with 1 mL MS-grade H2O a constant amount of 300 ug beads was added to each sample well. Protein precipitation was induced by the addition of acetonitrile to a final volume of 70%. To ensure complete precipitation, we incubated the suspension for 10 min at room temperature and 1200 rpm and allowed beads to settle down for further 10 min without agitation. Subsequent to the removal of supernatant on a magnetic rack (e.g. DynaMag™-96 Side Skirted Magnet, Prod. No. 12027, Invitrogen, Thermo Fisher Scientific, Darmstadt, Germany), beads were washed trice in 100% isopropanol for 10 min at 50°C and 1400 rpm to clear all paraffin. For the enzymatic digest, we estimated the protein yield from 25 mm2 starting material based on a previous evaluation for the processing for FFPE tissue (Coscia et al., 2020). Consequently, beads of each sample well were resuspended in 100 μL of 100 mM Tris, pH 8.5, supplemented with 0.5 μg of trypsin and LysC, respectively, and incubated overnight at 37°C and 1300 rpm. On the next day, we enclosed a post-digest of 4 hours at 37°C and 1300 rpm with 0.5 μg of both enzymes. Hereafter, the supernatant was removed completely while placing the 96-well plate on the magnetic rack and transferred twice to a 96-well PCR plate (twin.tec® PCR Plate LoBind®, semi-skirted, 250 μL, Prod. No. 0030129504, Eppendorf, Hamburg, Germany) to remove residual magnetic particles. The enzymatic reaction was quenched using TFA at a final concentration of 1% (v/v) and resulting peptides were stored at −20°C until further processing. To generate deep proteomic profiles for specific tissues such as the lung and the lymph nodes, 30-50 μg of peptides were pooled for the respective organ as well as separately for either samples derived from patients with SARS-CoV-2 or their controls. Each sample pool, herein called ‘libraries’, was subsequently fractionated into 24 fractions by high-pH reversed-phase chromatography using the ‘Spider fractionator’ as described previously (Kulak et al, 2017). In preparation for the LC-MS measurement using an Evosep LC system, peptides of the single-shot samples and the deep proteomes were loaded on C18 EvoTips according to the manufacturer’s instructions (Evosep, Odense, Denmark) and stored at 4°C until further processing.

The AssayMAP Bravo robot (Agilent) performed the enrichment for phosphopeptides (80 μg) by priming AssayMAP cartridges (packed with 5 μl Fe3+-NTA) with 0.1% TFA in 99% ACN followed by equilibration in equilibration buffer (1% TFA/80% ACN) and loading of the same amount of peptides resuspended in equilibration buffer. Enriched phosphopeptides were eluted with 1% ammonium hydroxide, which was evaporated using a Speedvac for 20 min. Dried peptides were resuspended in 6 μl 2% acetonitrile (v/v)/ 0.1% trifluoroacetic acid (v/v) and 5 μl were analyzed by LC–MS/MS

### Liquid chromatography and mass spectrometry

For the LC-MS/MS analyses of the full proteome, we coupled the Evosep One platform online to an Orbitrap Exploris 480 Mass Spectrometer (Thermo Fisher Scientific, Waltham, USA) using a nano-electrospray ion source (Thermo Fisher Scientific). Peptides were loaded on a 15 cm HPLC column (inner diameter: 150 μm; generated in-house with ReproSil-Pur C18-AQ 1.9 μm silica beads (Dr. Maisch GmbH, Ammerbuch, Germany)(Müller-Reif et al., 2021) that was kept at 60°C by an oven containing a Peltier element (in-house development). Each sample was separated on the analytical column employing the standardized gradient of 88 min (15 samples per day) from the Evosep+ method set. Data for single-shot samples were acquired in a data-independent (DIA) mode with full MS scans (scan range: 300 to 1,650 m/z; resolution: 120,000; maximum injection time: 50 ms; normalized AGC target: 300%) and 40 periodical MS/MS segments applying isolation windows based on peptide prediction in MaxQuant.live (resolution: 30,000; maximum injection time: 50 ms; normalized AGC target: 1000%). Peptide fragmentation was enabled using a normalized collision energy of 30%. Fractions of the pre-fractionated library were processed in data-dependent acquisition (DDA) mode (Top12 method) with full MS (scan range: 350 to 1,400 m/z; resolution: 60,000; maximum injection time: 25 ms; normalized AGC target: 300%) and MS/MS scans (resolution: 15,000, isolation window: 1.3 m/z, normalized collision energy: 30%, maximum injection time: 22 ms, normalized AGC target: 200%, dynamic exclusion: 30 sec). All spectra were acquired in profile mode using positive polarity.

For phospho proteomic analyses, samples were loaded onto a 50-cm reversed-phase column (75 μm inner diameter, packed in house with ReproSil-Pur C18-AQ 1.9 μm resin (Dr Maisch)). The column temperature was maintained at 60 °C using a homemade column oven. A binary buffer system, consisting of buffer A (0.1% FA) and buffer B (80% ACN plus 0.1% FA) was used for peptide separation, at a flow rate of 300 nl min−1. An EASY-nLC 1200 system (Thermo Fisher Scientific) for nano-flow liquid chromatography was directly coupled online with the mass spectrometer (Orbitrap Exploris 480, Thermo Fisher Scientific) via a nano-electrospray source. The FAIMS device was placed between the nanoelectrospray source and the mass spectrometer for spectral library generation. Spray voltage was set to 2,650 V, RF level to 40 and heated capillary temperature to 275 °C. 5 μl of phosphopeptide samples were loaded and eluted with a 70-min gradient starting at 3% buffer B followed by a stepwise increase to 19% in 40 min, 41% in 20 min, 90% in 5 min and 95% in 5 min. The mass spectrometer was operated in DIA mode (in profile mode using positive polarity) with a full scan range of 300– 1,400 m/z at 120,000 resolution at 200 m/z and a maximum fill time of 60 ms. One full scan was followed by 32 windows with a resolution of 30,000. Normalized AGC target and maximum fill time were set to 1,000% and 54 ms, respectively. Precursor ions were fragmented by HCD (NCE stepped 25–27.5–30%). For the library generation, pooled phospho-enriched sample was measured with 11 different CV settings (−30, −40, −45, −50, −55, −60, −65, −70, −75, −80 or −90 V) using the same DIA method. The noted single CVs were applied to the FAIMS electrodes throughout the analysis.

### Data processing

Single-shot data were processed organ-wise in Spectronaut version 14.7.201007.47784 (Copernicus, Biognosys AG, Schlieren, Switzerland) employing a library-free strategy (directDIA) and searching against the Uniprot human databases UP000005640_9606, UP000005640_9606_additional (integrated in the Spectronaut software; 21,039 and 70,579 entries, respectively) and the severe acute respiratory syndrome coronavirus 2 (downloaded on July 28 2020, 120 entries). Enzymatic cleavage was defined by Trypsin/P with a peptide length of 7-52 and a maximum number of two missed cleavages. Carbamidomethylation was set as a fixed modification while methionine oxidation and N-terminal acetylation were indicated as variable modifications. A minimum number of 3 and a maximum number of 6 were selected as Best N Fragment per peptides. Significance filtering followed a precursor and protein q-value cutoff of 1%. Other processing parameters were kept by standard settings of the Spectronaut version.

Spectronaut version 16.2.220903.53000 (Biognosys) (Copernicus, Biognosys AG, Schlieren, Switzerland) was used to generate DIA libraries and analyze DIA single-runs. The library consists out of DIA single-runs acquired with a static CV ranging from −30 to −90. For the generation of the hybrid library, the library runs were combined with cohort samples measured also as DIA single-runs, and phosphorylation at Serine/Threonine/Tyrosine was added as a variable modification to default settings. The maximum number of fragment ions per peptide was increased from 3 to 25. The biological replicate files were analyzed against the hybrid library. Phosphorylation at Serine/Threonine/Tyrosine was added as a variable modification to default settings with a disabled PTM localization filter. An FDR of 1% determined the significance level.

### Bioinformatics data analysis

Bioinformatics data analysis was performed using the R statistical computing environment version 4.0.2. In preparation for the analysis, protein intensities were log2-transformed. For the Principal Component Analysis (PCA) and ANOVA, protein identifications were filtered for a minimum of 80% valid values in each group and imputed sample-wise based on a normal distribution (width of 0.3, downshift of 1.8) (FactoMineR package). For the analysis of variance (ANOVA) testing, the R stats package was employed. P-values were adjusted using the Benjamini-Hochberg method, followed by post-hoc computation of the Tukey Honest Significant Differences (Tukey HSD) at a confidence level of 0.95. Differential expression was determined for proteins with a minimal identification of three valid values in total using Student’s t-tests (unpaired, variances assumed to be equal), p-value correction (qvalue package) and filtering for significant differential expression (q-value < 0.05, fold change >= 1.5). For the comparison of differential expression across organs, we calculated the ranking factor for each protein, defined as the product of fold-change and statistical consistency (q-value) (Xiao et al., 2014).

Marker proteins for coagulation, platelets and red blood cells (RBC) were extracted from (Karayel et al., 2022), the 30 most abundant and non-redundant proteins from the plasma were added from (Geyer et al., 2016). Proteins of this systemic-inflammatory response were complemented by immunoglobulins identified in our dataset (Supplementary Table 3). For the ‘filtering’ of systemic-inflammatory signatures, statistical tests were performed on the entire set of proteins and subsequently filtered based on the previously generated table.

Pearson correlations between groups were determined using the R stats package on median protein intensities per group only considering complete observations. Biological pathway enrichment of significantly differentially regulated proteins in each organ was performed for the entire dataset including the systemic-inflammatory and the organ-specific effects, respectively, using the WebGestalt gene set analysis toolkit for a GSEA (organism: homo sapiens, FDR threshold: 0.05, databases: ‘pathway_KEGG’, ‘pathway_Reactome’, ‘geneontology_Biological_ Process_noRedundant’). Boxplots, point-size, bar, radar and scatter (Volcano) plots were generated using the ggplot2 package, upset plots and heatmaps were generated via the ComplexHeatmap package.

To manually validate the quality of identified SARS-CoV-2 viral peptides, the corresponding fragment intensities were predicted using AlphaPeptDeep (Zeng et al., 2022) at normalized collisional energy 30, and were plotted with AlphaViz (Voytik et al., 2022), comparing them against the detected intensities at the apex elution positions. The elution profiles of the fragments in DIA data were extracted and plotted using AlphaViz. The matching mass error was set to 20 ppm for MS2 spectra.

For the analysis of the phospho-proteome, the quantification of phosphorylated peptides from the Spectronaut output table were collapsed using the plug-in tool for Perseus (1.6.7.0) to annotate the exact position of the phospho sites. For this, phospho sites were aggregated using the linear model-based approach and filtered for a localization probability > 0.75. The data were filtered to contain > 25% valid values across all samples. Raw intensities were log2 transformed and missing values were imputed based on a Gaussian normal distribution with a width of 0.3 and a downshift of 1.8. Overrepresentation analysis (ORA) of annotation terms was performed using the gprofiler2 package (organism: homo sapiens, user threshold: 0.05, databases: Reactome). Network representation of proteins with regulated sites was performed with the STRING app (1.5.1) in Cytoscape (3.7.2).

## References

Aebersold, R., and Mann, M. (2016). Mass-spectrometric exploration of proteome structure and function. Nature 537, 347–355.

Bache, N., Geyer, P.E., Bekker-Jensen, D.B., Hoerning, O., Falkenby, L., Treit, P.V., Doll, S., Paron, I., Muller, J.B., Meier, F., et al. (2018). A Novel LC System Embeds Analytes in Pre-formed Gradients for Rapid, Ultra-robust Proteomics. Mol Cell Proteomics 17, 2284–2296.

Badrani, J.H., Strohm, A.N., Lacasa, L., Civello, B., Cavagnero, K., Haung, Y.-A., Amadeo, M., Naji, L.H., Lund, S.J., Leng, A., et al. (2022). RNA-binding protein RBM3 intrinsically suppresses lung innate lymphoid cell activation and inflammation partially through CysLT1R. Nature Communications 13, 4435.

Batth, T.S., Tollenaere, M.X., Ruther, P., Gonzalez-Franquesa, A., Prabhakar, B.S., Bekker-Jensen, S., Deshmukh, A.S., and Olsen, J.V. (2019). Protein Aggregation Capture on Microparticles Enables Multipurpose Proteomics Sample Preparation. Mol Cell Proteomics 18, 1027–1035.

Bian, Y., Zheng, R., Bayer, F.P., Wong, C., Chang, Y.C., Meng, C., Zolg, D.P., Reinecke, M., Zecha, J., Wiechmann, S., et al. (2020). Robust, reproducible and quantitative analysis of thousands of proteomes by micro-flow LC-MS/MS. Nat Commun 11, 157.

Bojkova, D., Klann, K., Koch, B., Widera, M., Krause, D., Ciesek, S., Cinatl, J., and Munch, C. (2020). Proteomics of SARS-CoV-2-infected host cells reveals therapy targets. Nature 583, 469–472.

Bouhaddou, M., Memon, D., Meyer, B., White, K.M., Rezelj, V.V., Correa Marrero, M., Polacco, B.J., Melnyk, J.E., Ulferts, S., Kaake, R.M., et al. (2020). The Global Phosphorylation Landscape of SARS-CoV-2 Infection. Cell 182, 685–712 e619.

Camper, N., Glasgow, A.M., Osbourn, M., Quinn, D.J., Small, D.M., McLean, D.T., Lundy, F.T., Elborn, J.S., McNally, P., Ingram, R.J., et al. (2016). A secretory leukocyte protease inhibitor variant with improved activity against lung infection. Mucosal Immunol 9, 669–676.

COMBAT (2022). A blood atlas of COVID-19 defines hallmarks of disease severity and specificity. Cell 185, 916–938 e958.

Coscia, F., Doll, S., Bech, J.M., Schweizer, L., Mund, A., Lengyel, E., Lindebjerg, J., Madsen, G.I., Moreira, J.M., and Mann, M. (2020). A streamlined mass spectrometry-based proteomics workflow for large-scale FFPE tissue analysis. J Pathol 251, 100–112.

Daamen, A.R., Bachali, P., Owen, K.A., Kingsmore, K.M., Hubbard, E.L., Labonte, A.C., Robl, R., Shrotri, S., Grammer, A.C., and Lipsky, P.E. (2021). Comprehensive transcriptomic analysis of COVID-19 blood, lung, and airway. Sci Rep 11, 7052.

Daniloski, Z., Jordan, T.X., Wessels, H.H., Hoagland, D.A., Kasela, S., Legut, M., Maniatis, S., Mimitou, E.P., Lu, L., Geller, E., et al. (2021). Identification of Required Host Factors for SARS-CoV-2 Infection in Human Cells. Cell 184, 92–105 e116.

Douaud, G., Lee, S., Alfaro-Almagro, F., Arthofer, C., Wang, C.Y., McCarthy, P., Lange, F., Andersson, J.L.R., Griffanti, L., Duff, E., et al. (2022). SARS-CoV-2 is associated with changes in brain structure in UK Biobank. Nature 604, 697-+.

Durovic, E., Bien, C., Bien, C.G., and Isenmann, S. (2021). MOG antibody-associated encephalitis secondary to Covid-19: case report. BMC Neurol 21, 414.

Farber, D.L. (2021). Tissues, not blood, are where immune cells function. Nature 593, 506–509.

Flynn, R.A., Belk, J.A., Qi, Y., Yasumoto, Y., Wei, J., Alfajaro, M.M., Shi, Q., Mumbach, M.R., Limaye, A., DeWeirdt, P.C., et al. (2021). Discovery and functional interrogation of SARS-CoV-2 RNA-host protein interactions. Cell 184, 2394–2411 e2316.

Foll, M.C., Fahrner, M., Oria, V.O., Kuhs, M., Biniossek, M.L., Werner, M., Bronsert, P., and Schilling, O. (2018). Reproducible proteomics sample preparation for single FFPE tissue slices using acid-labile surfactant and direct trypsinization. Clin Proteomics 15, 11.

Frieman, M., Yount, B., Heise, M., Kopecky-Bromberg, S.A., Palese, P., and Baric, R.S. (2007). Severe acute respiratory syndrome coronavirus ORF6 antagonizes STAT1 function by sequestering nuclear import factors on the rough endoplasmic reticulum/Golgi membrane. J Virol 81, 9812–9824.

Gao, H., Liu, Y., Demichev, V., Tate, S., Chen, C., Zhu, J., Lu, C., Ralser, M., Guo, T., and Zhu, Y. (2022). Optimization of Microflow LC Coupled with Scanning SWATH and Its Application in Hepatocellular Carcinoma Tissues. J Proteome Res 21, 1686–1693.

Geyer, P.E., Arend, F.M., Doll, S., Louiset, M.L., Virreira Winter, S., Muller-Reif, J.B., Torun, F.M., Weigand, M., Eichhorn, P., Bruegel, M., et al. (2021). High-resolution serum proteome trajectories in COVID-19 reveal patient-specific seroconversion. EMBO Mol Med 13, e14167.

Geyer, P.E., Kulak, N.A., Pichler, G., Holdt, L.M., Teupser, D., and Mann, M. (2016). Plasma Proteome Profiling to Assess Human Health and Disease. Cell Syst 2, 185–195.

Geyer, P.E., Voytik, E., Treit, P.V., Doll, S., Kleinhempel, A., Niu, L., Muller, J.B., Buchholtz, M.L., Bader, J.M., Teupser, D., et al. (2019). Plasma Proteome Profiling to detect and avoid sample-related biases in biomarker studies. EMBO Mol Med 11, e10427.

Gillespie, M., Jassal, B., Stephan, R., Milacic, M., Rothfels, K., Senff-Ribeiro, A., Griss, J., Sevilla, C., Matthews, L., Gong, C., et al. (2022). The reactome pathway knowledgebase 2022. Nucleic Acids Res 50, D687–D692.

Gordon, D.E., Jang, G.M., Bouhaddou, M., Xu, J., Obernier, K., White, K.M., O’Meara, M.J., Rezelj, V.V., Guo, J.Z., Swaney, D.L., et al. (2020). A SARS-CoV-2 protein interaction map reveals targets for drug repurposing. Nature 583, 459–468.

Guan, Y., Guo, W., Zou, S.P., Dubner, R., and Ren, K. (2003). Inflammation-induced upregulation of AMPA receptor subunit expression in brain stem pain modulatory circuitry. Pain 104, 401–413.

Hirschbühl, K., Dintner, S., Beer, M., Wylezich, C., Schlegel, J., Delbridge, C., Borcherding, L., Lippert, J., Schiele, S., Muller, G., et al. (2021). Viral mapping in COVID-19 deceased in the Augsburg autopsy series of the first wave: A multiorgan and multimethodological approach. PLoS One 16, e0254872.

Huang, C., Wang, Y., Li, X., Ren, L., Zhao, J., Hu, Y., Zhang, L., Fan, G., Xu, J., Gu, X., et al. (2020). Clinical features of patients infected with 2019 novel coronavirus in Wuhan, China. Lancet 395, 497–506.

Hughes, C.S., Moggridge, S., Muller, T., Sorensen, P.H., Morin, G.B., and Krijgsveld, J. (2019). Single-pot, solid-phase-enhanced sample preparation for proteomics experiments. Nat Protoc 14, 68–85.

Initiative, C.-H.G. (2021). Mapping the human genetic architecture of COVID-19. Nature 600, 472–477.

Jackson, C.B., Farzan, M., Chen, B., and Choe, H. (2022). Mechanisms of SARS-CoV-2 entry into cells. Nat Rev Mol Cell Biol 23, 3–20.

Jiang, X., Jiang, X., Feng, S., Tian, R., Ye, M., and Zou, H. (2007). Development of efficient protein extraction methods for shotgun proteome analysis of formalin-fixed tissues. J Proteome Res 6, 1038–1047.

Johnsson, M., Asztely, F., Hejnebo, S., Axelsson, M., Malmestrom, C., Olausson, T., and Lycke, J. (2022). SARS-COV-2 a trigger of myelin oligodendrocyte glycoprotein-associated disorder. Ann Clin Transl Neurol 9, 1296–1301.

Kanehisa, M., Furumichi, M., Sato, Y., Kawashima, M., and Ishiguro-Watanabe, M. (2022). KEGG for taxonomy-based analysis of pathways and genomes. Nucleic Acids Res.

Karayel, O., Virreira Winter, S., Padmanabhan, S., Kuras, Y.I., Vu, D.T., Tuncali, I., Merchant, K., Wills, A.M., Scherzer, C.R., and Mann, M. (2022). Proteome profiling of cerebrospinal fluid reveals biomarker candidates for Parkinson’s disease. Cell Rep Med 3, 100661.

Kaya, T., Mattugini, N., Liu, L., Ji, H., Cantuti-Castelvetri, L., Wu, J., Schifferer, M., Groh, J., Martini, R., Besson-Girard, S., et al. (2022). CD8(+) T cells induce interferon-responsive oligodendrocytes and microglia in white matter aging. Nat Neurosci 25, 1446–1457.

Kim, D., Kim, S., Park, J., Chang, H.R., Chang, J., Ahn, J., Park, H., Park, J., Son, N., Kang, G., et al. (2021a). A high-resolution temporal atlas of the SARS-CoV-2 translatome and transcriptome. Nat Commun 12, 5120.

Kim, S.G., Lee, S., Kim, Y., Park, J., Woo, D., Kim, D., Li, Y., Shin, W., Kang, H., Yook, C., et al. (2021b). Tanc2-mediated mTOR inhibition balances mTORC1/2 signaling in the developing mouse brain and human neurons. Nature Communications 12.

Kristensen, A.S., Jenkins, M.A., Banke, T.G., Schousboe, A., Makino, Y., Johnson, R.C., Huganir, R., and Traynelis, S.F. (2011). Mechanism of Ca2+/calmodulin-dependent kinase II regulation of AMPA receptor gating. Nat Neurosci 14, 727–735.

Lakshmikanthan, S., Zieba, B.J., Ge, Z.D., Momotani, K., Zheng, X., Lund, H., Artamonov, M.V., Maas, J.E., Szabo, A., Zhang, D.X., et al. (2014). Rap1b in smooth muscle and endothelium is required for maintenance of vascular tone and normal blood pressure. Arterioscler Thromb Vasc Biol 34, 1486–1494.

Lu, R., Zhao, X., Li, J., Niu, P., Yang, B., Wu, H., Wang, W., Song, H., Huang, B., Zhu, N., et al. (2020). Genomic characterisation and epidemiology of 2019 novel coronavirus: implications for virus origins and receptor binding. Lancet 395, 565–574.

Luo, H., Liu, D., Liu, W., Wang, G., Chen, L., Cao, Y., Wei, J., Xiao, M., Liu, X., Huang, G., et al. (2021). Germline variants in UNC13D and AP3B1 are enriched in COVID-19 patients experiencing severe cytokine storms. Eur J Hum Genet 29, 1312–1315.

Mao, R., Qiu, Y., He, J.S., Tan, J.Y., Li, X.H., Liang, J., Shen, J., Zhu, L.R., Chen, Y., Iacucci, M., et al. (2020). Manifestations and prognosis of gastrointestinal and liver involvement in patients with COVID-19: a systematic review and meta-analysis. Lancet Gastroenterol Hepatol 5, 667–678.

Martinez-Martinez, E., Ibarrola, J., Fernandez-Celis, A., Santamaria, E., Fernandez-Irigoyen, J., Rossignol, P., Jaisser, F., and Lopez-Andres, N. (2017). Differential Proteomics Identifies Reticulocalbin-3 as a Novel Negative Mediator of Collagen Production in Human Cardiac Fibroblasts. Sci Rep 7, 12192.

Matschke, J., Lutgehetmann, M., Hagel, C., Sperhake, J.P., Schroder, A.S., Edler, C., Mushumba, H., Fitzek, A., Allweiss, L., Dandri, M., et al. (2020). Neuropathology of patients with COVID-19 in Germany: a post-mortem case series. Lancet Neurol 19, 919–929.

Mavrikaki, M., Lee, J.D., Solomon, I.H., and Slack, F.J. (2022). Severe COVID-19 is associated with molecular signatures of aging in the human brain. Nature Aging.

Mecham, R.P. (2018). Elastin in lung development and disease pathogenesis. Matrix Biol 73, 6–20.

Melms, J.C., Biermann, J., Huang, H., Wang, Y., Nair, A., Tagore, S., Katsyv, I., Rendeiro, A.F., Amin, A.D., Schapiro, D., et al. (2021). A molecular single-cell lung atlas of lethal COVID-19. Nature 595, 114–119.

Messner, C.B., Demichev, V., Bloomfield, N., Yu, J.S.L., White, M., Kreidl, M., Egger, A.S., Freiwald, A., Ivosev, G., Wasim, F., et al. (2021). Ultra-fast proteomics with Scanning SWATH. Nat Biotechnol 39, 846–854.

Messner, C.B., Demichev, V., Wendisch, D., Michalick, L., White, M., Freiwald, A., Textoris-Taube, K., Vernardis, S.I., Egger, A.S., Kreidl, M., et al. (2020). Ultra-High-Throughput Clinical Proteomics Reveals Classifiers of COVID-19 Infection. Cell Syst 11, 11–24 e14.

Mi, H., Muruganujan, A., Ebert, D., Huang, X., and Thomas, P.D. (2019). PANTHER version 14: more genomes, a new PANTHER GO-slim and improvements in enrichment analysis tools. Nucleic Acids Res 47, D419–D426.

Miorin, L., Kehrer, T., Sanchez-Aparicio, M.T., Zhang, K., Cohen, P., Patel, R.S., Cupic, A., Makio, T., Mei, M., Moreno, E., et al. (2020). SARS-CoV-2 Orf6 hijacks Nup98 to block STAT nuclear import and antagonize interferon signaling. Proc Natl Acad Sci U S A 117, 28344–28354.

Miyamoto, Y., Itoh, Y., Suzuki, T., Tanaka, T., Sakai, Y., Koido, M., Hata, C., Wang, C.X., Otani, M., Moriishi, K., et al. (2022). SARS-CoV-2 ORF6 disrupts nucleocytoplasmic trafficking to advance viral replication. Commun Biol 5, 483.

Müller-Reif, J.B., Hansen, F.M., Schweizer, L., Treit, P.V., Geyer, P.E., and Mann, M. (2021). A New Parallel High-Pressure Packing System Enables Rapid Multiplexed Production of Capillary Columns. Mol Cell Proteomics 20, 100082.

Muller, T., Kalxdorf, M., Longuespee, R., Kazdal, D.N., Stenzinger, A., and Krijgsveld, J. (2020). Automated sample preparation with SP3 for low-input clinical proteomics. Mol Syst Biol 16, e9111.

Namkoong, H., Edahiro, R., Takano, T., Nishihara, H., Shirai, Y., Sonehara, K., Tanaka, H., Azekawa, S., Mikami, Y., Lee, H., et al. (2022). DOCK2 is involved in the host genetics and biology of severe COVID-19. Nature 609, 754–760.

Nie, X., Qian, L., Sun, R., Huang, B., Dong, X., Xiao, Q., Zhang, Q., Lu, T., Yue, L., Chen, S., et al. (2021). Multi-organ proteomic landscape of COVID-19 autopsies. Cell 184, 775–791 e714.

Overmyer, K.A., Shishkova, E., Miller, I.J., Balnis, J., Bernstein, M.N., Peters-Clarke, T.M., Meyer, J.G., Quan, Q., Muehlbauer, L.K., Trujillo, E.A., et al. (2021). Large-Scale Multi-omic Analysis of COVID-19 Severity. Cell Syst 12, 23–40 e27.

Pairo-Castineira, E., Clohisey, S., Klaric, L., Bretherick, A.D., Rawlik, K., Pasko, D., Walker, S., Parkinson, N., Fourman, M.H., Russell, C.D., et al. (2021). Genetic mechanisms of critical illness in COVID-19. Nature 591, 92–98.

Quintero-Rivera, F., Xi, Q.J., Keppler-Noreuil, K.M., Lee, J.H., Higgins, A.W., Anchan, R.M., Roberts, A.E., Seong, I.S., Fan, X., Lage, K., et al. (2015). MATR3 disruption in human and mouse associated with bicuspid aortic valve, aortic coarctation and patent ductus arteriosus. Hum Mol Genet 24, 2375–2389.

Rubin, E.J., Longo, D.L., and Baden, L.R. (2021). Interleukin-6 Receptor Inhibition in Covid-19 - Cooling the Inflammatory Soup. N Engl J Med 384, 1564–1565.

Sadarangani, M., Marchant, A., and Kollmann, T.R. (2021). Immunological mechanisms of vaccine-induced protection against COVID-19 in humans. Nat Rev Immunol 21, 475–484.

Safaiyan, S., Kannaiyan, N., Snaidero, N., Brioschi, S., Biber, K., Yona, S., Edinger, A.L., Jung, S., Rossner, M.J., and Simons, M. (2016). Age-related myelin degradation burdens the clearance function of microglia during aging. Nat Neurosci 19, 995–998.

Schaller, T., Hirschbuhl, K., Burkhardt, K., Braun, G., Trepel, M., Markl, B., and Claus, R. (2020). Postmortem Examination of Patients With COVID-19. JAMA 323, 2518–2520.

Schwabenland, M., Salie, H., Tanevski, J., Killmer, S., Lago, M.S., Schlaak, A.E., Mayer, L., Matschke, J., Puschel, K., Fitzek, A., et al. (2021). Deep spatial profiling of human COVID-19 brains reveals neuroinflammation with distinct microanatomical microglia-T-cell interactions. Immunity 54, 1594-+.

Shao, S., Fang, H., Duan, L., Ye, X., Rao, S., Han, J., Li, Y., Yuan, G., Liu, W., and Zhang, X. (2020). Lysyl hydroxylase 3 increases collagen deposition and promotes pulmonary fibrosis by activating TGFbeta1/Smad3 and Wnt/beta-catenin pathways. Arch Med Sci 16, 436–445.

Stephenson, E., Reynolds, G., Botting, R.A., Calero-Nieto, F.J., Morgan, M.D., Tuong, Z.K., Bach, K., Sungnak, W., Worlock, K.B., Yoshida, M., et al. (2021). Single-cell multi-omics analysis of the immune response in COVID-19. Nat Med 27, 904–916.

Stukalov, A., Girault, V., Grass, V., Karayel, O., Bergant, V., Urban, C., Haas, D.A., Huang, Y., Oubraham, L., Wang, A., et al. (2021). Multilevel proteomics reveals host perturbations by SARS-CoV-2 and SARS-CoV. Nature 594, 246–252.

Tjan, L.H., Furukawa, K., Nagano, T., Kiriu, T., Nishimura, M., Arii, J., Hino, Y., Iwata, S., Nishimura, Y., and Mori, Y. (2021). Early Differences in Cytokine Production by Severity of Coronavirus Disease 2019. J Infect Dis 223, 1145–1149.

Varga, Z., Flammer, A.J., Steiger, P., Haberecker, M., Andermatt, R., Zinkernagel, A.S., Mehra, M.R., Schuepbach, R.A., Ruschitzka, F., and Moch, H. (2020). Endothelial cell infection and endotheliitis in COVID-19. Lancet 395, 1417–1418.

Venkataramani, V., and Winkler, F. (2022). Cognitive Deficits in Long Covid-19. N Engl J Med 387, 1813–1815.

Voytik, E., Skowronek, P., Zeng, W.-F., Tanzer, M.C., Brunner, A.-D., Thielert, M., Strauss, M.T., Willems, S., and Mann, M. (2022). AlphaViz: Visualization and validation of critical proteomics data directly at the raw data level. bioRxiv, 2022.2007.2012.499676.

Weldon, S., McNally, P., McElvaney, N.G., Elborn, J.S., McAuley, D.F., Wartelle, J., Belaaouaj, A., Levine, R.L., and Taggart, C.C. (2009). Decreased levels of secretory leucoprotease inhibitor in the Pseudomonas-infected cystic fibrosis lung are due to neutrophil elastase degradation. J Immunol 183, 8148–8156.

Wisniewski, J.R., Dus, K., and Mann, M. (2013). Proteomic workflow for analysis of archival formalin-fixed and paraffin-embedded clinical samples to a depth of 10 000 proteins. Proteomics Clin Appl 7, 225–233.

Wojcechowskyj, J.A., Didigu, C.A., Lee, J.Y., Parrish, N.F., Sinha, R., Hahn, B.H., Bushman, F.D., Jensen, S.T., Seeholzer, S.H., and Doms, R.W. (2013). Quantitative phosphoproteomics reveals extensive cellular reprogramming during HIV-1 entry. Cell Host Microbe 13, 613–623.

Xiao, Y., Hsiao, T.H., Suresh, U., Chen, H.I., Wu, X., Wolf, S.E., and Chen, Y. (2014). A novel significance score for gene selection and ranking. Bioinformatics 30, 801–807.

Yalcin Kehribar, D., Cihangiroglu, M., Sehmen, E., Avci, B., Capraz, A., Yildirim Bilgin, A., Gunaydin, C., and Ozgen, M. (2021). The receptor for advanced glycation end product (RAGE) pathway in COVID-19. Biomarkers 26, 114–118.

Zeng, W.F., Zhou, X.X., Willems, S., Ammar, C., Wahle, M., Bludau, I., Voytik, E., Strauss, M.T., and Mann, M. (2022). AlphaPeptDeep: a modular deep learning framework to predict peptide properties for proteomics. Nat Commun 13, 7238.

