## Supplementary Tables and Figures for "Quantitative multi-organ proteomics of fatal COVID-19 uncovers tissue-specific effects beyond inflammation"

#### Abstract

**SARS-CoV-2 directly damages lung tissue via its infection and replication process and indirectly due to systemic effects of the host immune system. There are few systems-wide, untargeted studies of these effects on the different tissues of the human body and nearly all of them base their conclusions on the transcriptome. Here we developed a parallelized mass spectrometry (MS)-based proteomics workflow allowing the rapid, quantitative analysis of hundreds of virus-infected and FFPE preserved tissues. The first layer of response in all tissues was dominated by circulating inflammatory molecules. To discriminated between these systemic and true tissue-specific effects, we developed an analysis pipeline revealing that proteome alterations reflect extensive tissue damage, mostly similar to non-COVID diffuse alveolar damage. The next most affected organs were kidney and liver, while the lymph-vessel system was also strongly affected. Finally, secondary inflammatory effects of the brain correlated with receptor rearrangements and the degradation of neuronal myelin. Our results establish MS-based tissue proteomics as a promising strategy to inform organ-specific therapeutic interventions following COVID-19 infections.**

### Supplementary Information

#### Supplementary Table 1: Basic clinical patient characteristics

| ***Variable*** | ***No.*** |
|---|---|
| **Total number of patients** | 19 (100%) |
| Mean/Median Age (range) | 74/73 years (57–90) |
| Sex (male/female) | 14 (74%) / 5 (26%) |
| Smoker (yes/no) | 7 (37%) /12 (63%) |
| **Comorbidities** | |
| Median number (range) | 4 (0–9) |
| **Cardiovascular** | 13 (68%) |
| Atrial fibrillation | 11 (58%) |
| Coronary artery disease | 5 (26%) |
| Cardiomyopathy | 5 (26%) |
| Aortic valve stenosis | 1 (5%) |
| Hypertension | 13 (68%) |
| Arteriosclerosis | 9 (47%) |
| **Metabolic** | 11 (58%) |
| Diabetes | 6 (32%) |
| Obesity | 9 (47%) |
| Median body mass index (range) | 28.3 kg/m² (19.6 – 66.2) |
| **Chronic respiratory disease** | 5 (26%) |
| **Chronic renal disease** | 7 (37%) |
| **Hyperlipoproteinemia** | 4 (21%) |
| **Prevalent malignancies** | 3 (16%) |
| **Therapeutics** | |
| Angiotensin converting enzyme inhibitors | 9 (47%) |
| Angiotensin II receptor blockers | 2 (11%) |

CLL = chronic lymphatic leukemia, CMML = chronic myelomonocytic leukemia

#### Supplementary Table 2: Clinical course and treatment

| ***Variable*** | ***No.*** |
|---|---|
| **COVID-19 testing: nasopharyngeal swab** | 19 (100%) |
| **Symptoms on admission** | |
| Cough | 13 (68%) |
| Dyspnea | 11 (58%) |
| Fever | 9 (47%) |
| Worsening general condition | 9 (47%) |
| Diarrhea | 1 (5%) |
| Median time between onset of symptoms and admission* (range) | 5 days (0 – 21) |
| **Radiologic findings (CT = 4, X-Ray = 15)** | |
| Bilateral patchy shadowing | 16 (84%) |
| Ground-glass opacity | 2 (11%) |
| **Respiratory findings** | |
| Median Horowitz index* (range)) | 107 mmHg (50 – 150) |
| Acute respiratory distress syndrome | 16 (84%) |
| **Septic shock** | 7 (37%) |
| **Acute kidney injury** | 14 (74%) |
| **Congestive heart failure** | 13 (68%) |
| **Clinical diagnosed pneumonia** | 8 (42%) |
| **Thromboembolic events (DVT/PE)** | 2 (11%) |
| **Systemic therapy** | |
| Systemic glucocorticoids (hydrocortisone for septic shock) | 3 (16%) |
| Remdesivir | 0 (0%) |
| Chloroquine or Hydroxychloroquine | 8 (42%) |
| Reconvalescent plasma | 3 (16%) |
| Intravenous antibiotics | 17 (89%) |
| Median number of antibiotic substances (range) | 2 (0 – 8) |
| **Anticoagulation use** | |
| Deep venous thrombosis prophylaxis | 9 (47%) |
| Full dose anticoagulation | 10 (53%) |
| Vasopressors | 10 (53%) |
| **Ventilation** | |
| Oxygen only | 5 (26%) |
| Noninvasive | 5 (26%) |

| | |
|---|---|
| Invasive | 9 (47%) |
| **Renal replacement therapy** | 5 (26%) |

*Missing information in three cases
CT = computed tomography, DVT = deep vein thrombosis, PE = pulmonary embolism

#### Supplementary Table 3: List of quality control markers

A. Tab Panel markers - to assess the contribution of the systemic-inflammatory response
B. Tab Systemic-inflammatory response - common proteins of the plasma proteome

#### Supplementary Table 4: Differential protein expression of COVID-19 vs. other lung pathologies

A. Tab Differential protein expression
B. Tab GSEA enrichment across lung diseases

#### Supplementary Table 5: Differential regulation of protein phosphorylation in the lungs

A. Tab Differential regulation phosphorylation

#### Supplementary Table 6: Differential protein expression of healthy and control samples in each organ

A. Tab Differential protein expression
B. Tab GSEA enrichment across organs

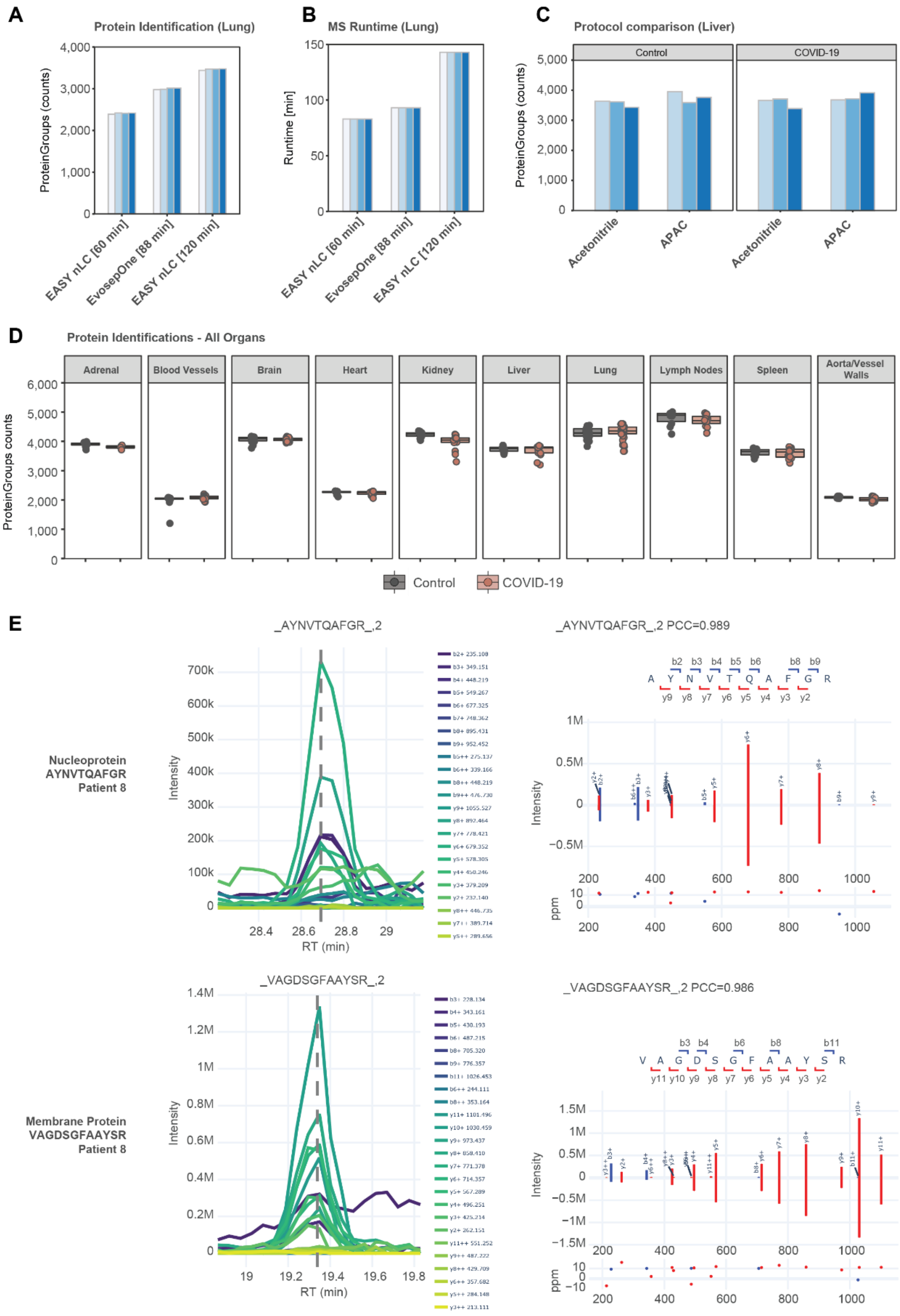
A
Protein Identification (Lung)
ProteinGroups (counts)
4,000
3,000
2,000
1,000
0
EASY nLC [60 min]
EvosepOne [88 min]
EASY nLC [120 min]
B
MS Runtime (Lung)
Runtime [min]
150
100
50
0
EASY nLC [60 min]
EvosepOne [88 min]
EASY nLC [120 min]
C
Protocol comparison (Liver)
Control
COVID-19
ProteinGroups (counts)
5,000
4,000
3,000
2,000
1,000
0
Acetonitrile
APAC
Acetonitrile
APAC
D
Protein Identifications - All Organs
Adrenal
Blood Vessels
Brain
Heart
Kidney
Liver
Lung
Lymph Nodes
Spleen
Aorta/Vessel Walls
ProteinGroups counts
6,000
5,000
4,000
3,000
2,000
1,000
0
Control
COVID-19
E
Nucleoprotein AYNVTQAFGR Patient 8
_AYNVTQAFGR_.2
Intensity
700k
600k
500k
400k
300k
200k
100k
0
28.4
28.6
28.8
29
RT (min)
_AYNVTQAFGR_.2 PCC=0.989
b2 b3 b4 b5 b6 b8 b9
A Y N V T Q A F G R
y9 y8 y7 y6 y5 y4 y3 y2
Intensity
1M
0.5M
0
−0.5M
200
400
600
800
1000
ppm
10
0
Membrane Protein VAGDSGFAAYSR Patient 8
_VAGDSGFAAYSR_.2
Intensity
1.4M
1.2M
1M
0.8M
0.6M
0.4M
0.2M
0
19
19.2
19.4
19.6
19.8
RT (min)
_VAGDSGFAAYSR_.2 PCC=0.986
b3 b4 b6 b8 b11
V A G D S G F A A Y S R
y11 y10 y9 y8 y7 y6 y5 y4 y3 y2
Intensity
1.5M
1M
0.5M
0
−0.5M
−1M
−1.5M
200
400
600
800
1000
ppm
10
0
−10

#### Supplementary Figure 1

**A** Comparison of protein identification in lung tissue of the COVID-19 cohort between the standardized 88 min gradient on Evosep One and a 60 min or 120 min gradient on the EASY nanoLC system, respectively.

**B** MS runtime of the comparison depicted in **B**.

**C** Evaluation of our workflow for streamlined deparaffinization and lysis (APAC) and our previous state-of-art protocol for the processing of deparaffinized FFPE tissue in a 96-well format (Coscia et al., 2020).

**D** Counts of quantified protein groups in COVID-19 samples and corresponding control groups in all organs using direct DIA data processing (not using a separately acquired DDA library).

**E** Extracted ion chromatograms and prediction of fragment intensities for two examples of SARS-CoV-2 peptides in the directDIA acquisition mode of the lungs. For both peptides, the same patient was selected to represent the cohort.

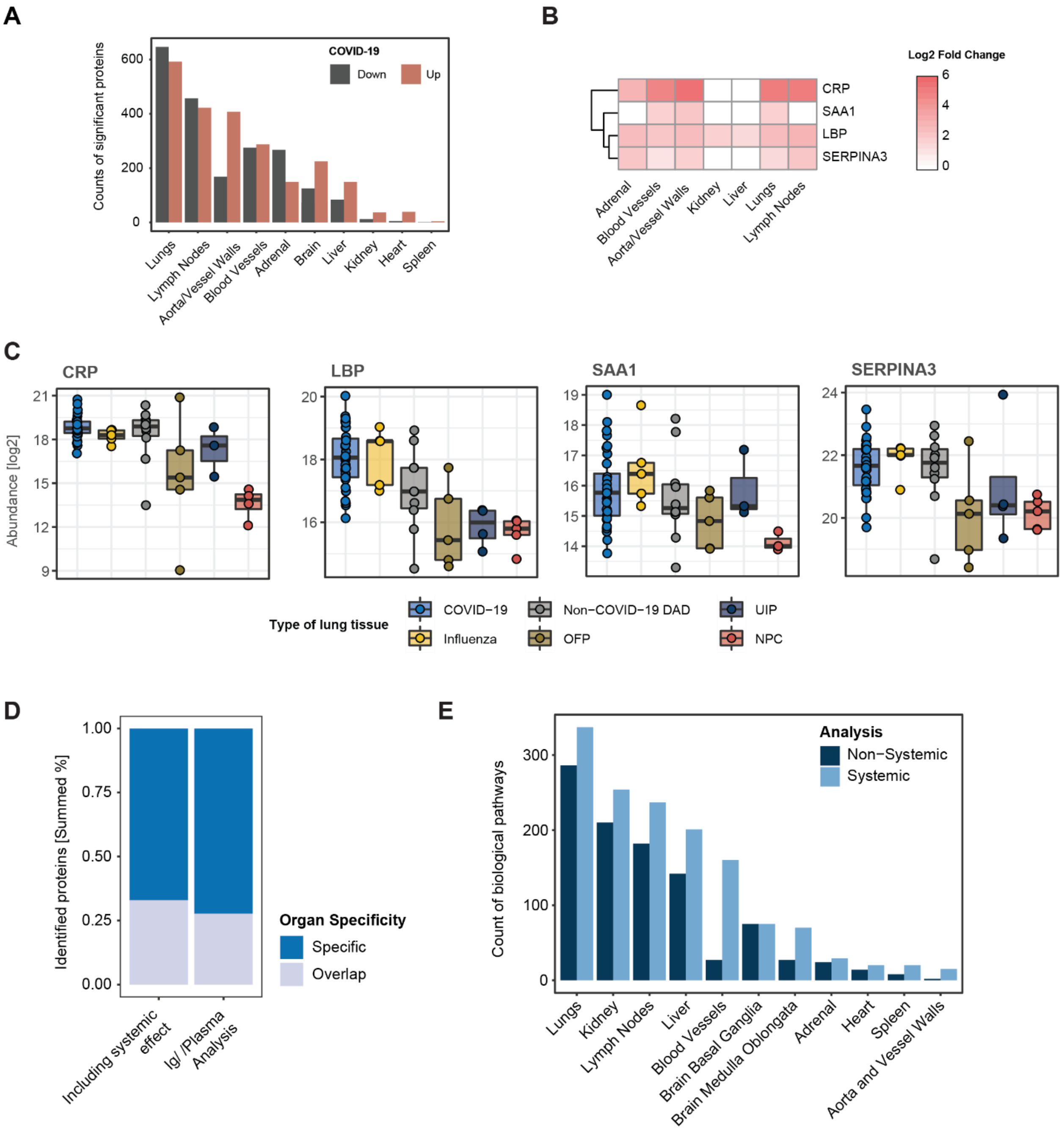
A
Counts of significant proteins
COVID-19
Down
Up
Lungs
Lymph Nodes
Aorta/Vessel Walls
Blood Vessels
Adrenal
Brain
Liver
Kidney
Heart
Spleen
B
Log2 Fold Change
CRP
SAA1
LBP
SERPINA3
Adrenal
Blood Vessels
Aorta/Vessel Walls
Kidney
Liver
Lungs
Lymph Nodes
C
CRP
LBP
SAA1
SERPINA3
Abundance [log2]
Type of lung tissue
COVID−19
Influenza
Non−COVID−19 DAD
OFP
UIP
NPC
D
Identified proteins [Summed %]
Including systemic effect
Ig/ /Plasma Analysis
Organ Specificity
Specific
Overlap
E
Count of biological pathways
Analysis
Non−Systemic
Systemic
Lungs
Kidney
Lymph Nodes
Liver
Blood Vessels
Brain Basal Ganglia
Brain Medulla Oblongata
Adrenal
Heart
Spleen
Aorta and Vessel Walls

#### Supplementary Figure 2

**A** Number of significantly differentially regulated proteins (COVID-19 vs controls) across all organs (minimum fold-change 1.5 at a q-value of 0.05).

**B** Fold changes (log2) of plasma-derived markers and predictive proteins for a severe progression of COVID-19.

**C** Boxplot representing the log2-abundance of the proteins shown in **B** for COVID-19 and similar pathologies of the lung which have been included into our study.

**D** Summed percentage of significantly differentially regulated proteins (minimum fold-change 1.5 at a q-value of 0.05) for the original proteome and after identification of the systemic effect, calculated for organ specificity and overlap in at least two organs, respectively.

**E** Count of biological pathways derived from an enrichment using the Reactome, KEGG and GO biological process databases and shown for differentially regulated proteins with association to the systemic and non-systemic subset of proteins across all organs.

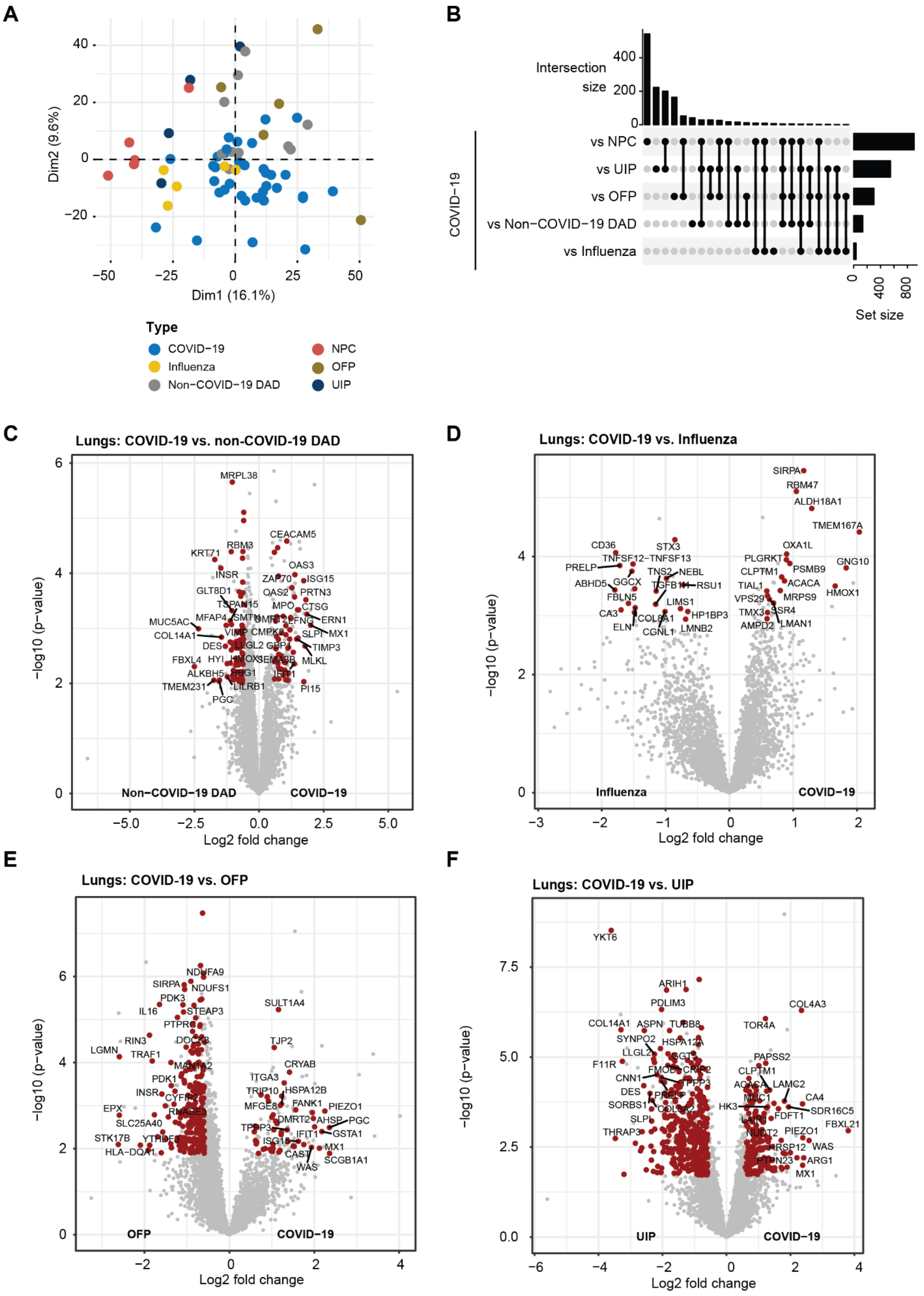
A
Dim2 (9.6%)
Dim1 (16.1%)
Type
COVID−19
Influenza
Non−COVID−19 DAD
NPC
OFP
UIP
B
Intersection size
COVID−19
vs NPC
vs UIP
vs OFP
vs Non−COVID−19 DAD
vs Influenza
Set size
C
Lungs: COVID-19 vs. non-COVID-19 DAD
−log10 (p−value)
Log2 fold change
Non−COVID−19 DAD
COVID−19
MRPL38
CEACAM5
RBM3
KRT71
OAS3
INSR
ZAP70
ISG15
GLT8D1
OAS2
PRTN3
TSPAN15
MPO
CTSG
MUC5AC
MFAP4
SMTN
DMRT2
LFNG
ERN1
COL14A1
VIMP
CMPK2
SLPI
MX1
DES
LLGL2
GBP1
TIMP3
FBXL4
HYI
HMOX1
SEMA3B
MLKL
ALKBH5
HBG1
IFIT1
TMEM231
LILRB1
PI15
PGC
D
Lungs: COVID-19 vs. Influenza
−log10 (p−value)
Log2 fold change
Influenza
COVID−19
SIRPA
RBM47
ALDH18A1
TMEM167A
CD36
STX3
OXA1L
TNFSF12−TNFSF13
PLGRKT
GNG10
PRELP
TNS2
NEBL
CLPTM1
PSMB9
ABHD5
GGCX
TGFB1I1
RSU1
TIAL1
ACACA
HMOX1
FBLN5
LIMS1
VPS29
MRPS9
CA3
COL8A1
HP1BP3
TMX3
SSR4
ELN
CGNL1
LMNB2
AMPD2
LMAN1
E
Lungs: COVID-19 vs. OFP
−log10 (p−value)
Log2 fold change
OFP
COVID−19
NDUFA9
SIRPA
NDUFS1
PDK3
SULT1A4
IL16
STEAP3
PTPRC
RIN3
DOCK8
TJP2
LGMN
TRAF1
MAN1A2
CRYAB
PDK1
ITGA3
INSR
CYFIP2
TRIP10
HSPA12B
EPX
RNASE3
MFGE8
FANK1
PIEZO1
SLC25A40
DMRT2
AHSP
PGC
TPPP3
GSTA1
STK17B
YTHDF3
ISG15
IFIT1
HLA−DQA1
CAST
MX1
WAS
SCGB1A1
F
Lungs: COVID-19 vs. UIP
−log10 (p−value)
Log2 fold change
UIP
COVID−19
YKT6
ARIH1
PDLIM3
COL4A3
COL14A1
ASPN
TUBB8
TOR4A
SYNPO2
HSPA12A
LLGL2
GGT5
PAPSS2
F11R
FMOD
CRIP2
CLPTM1
CNN1
TPPP3
ACACA
LAMC2
DES
PRELP
MUC1
CA4
SORBS1
COL6A2
HK3
SDR16C5
SLPI
FDFT1
FBXL21
THRAP3
NUDT2
PIEZO1
RSP12
WAS
PTPN23
ARG1
MX1

#### Supplementary Figure 3

**A** Principal component analysis (PCA) of lung samples derived from COVID-19 and lung pathologies sharing similar phenotype features as well as the non-pathological control group.

**B** Upset plot depicting the intersection of differentially regulated proteins between COVID-19 and lung pathologies sharing similar phenotype features as well as the non-pathological control group.

**C - F** Differential regulation of phosphor sites derived from pairwise comparisons between COVID-19 and the control groups of the lungs: non-COVID-19 DAD (**A**), Influenza (**B**), fibrosing organizing pneumonia (OFP, **C**) and usual interstitial pneumonia (UIP, **D**). Protein significance (t-test, q-val < 0.05, fold change >1.5) is highlighted in red.

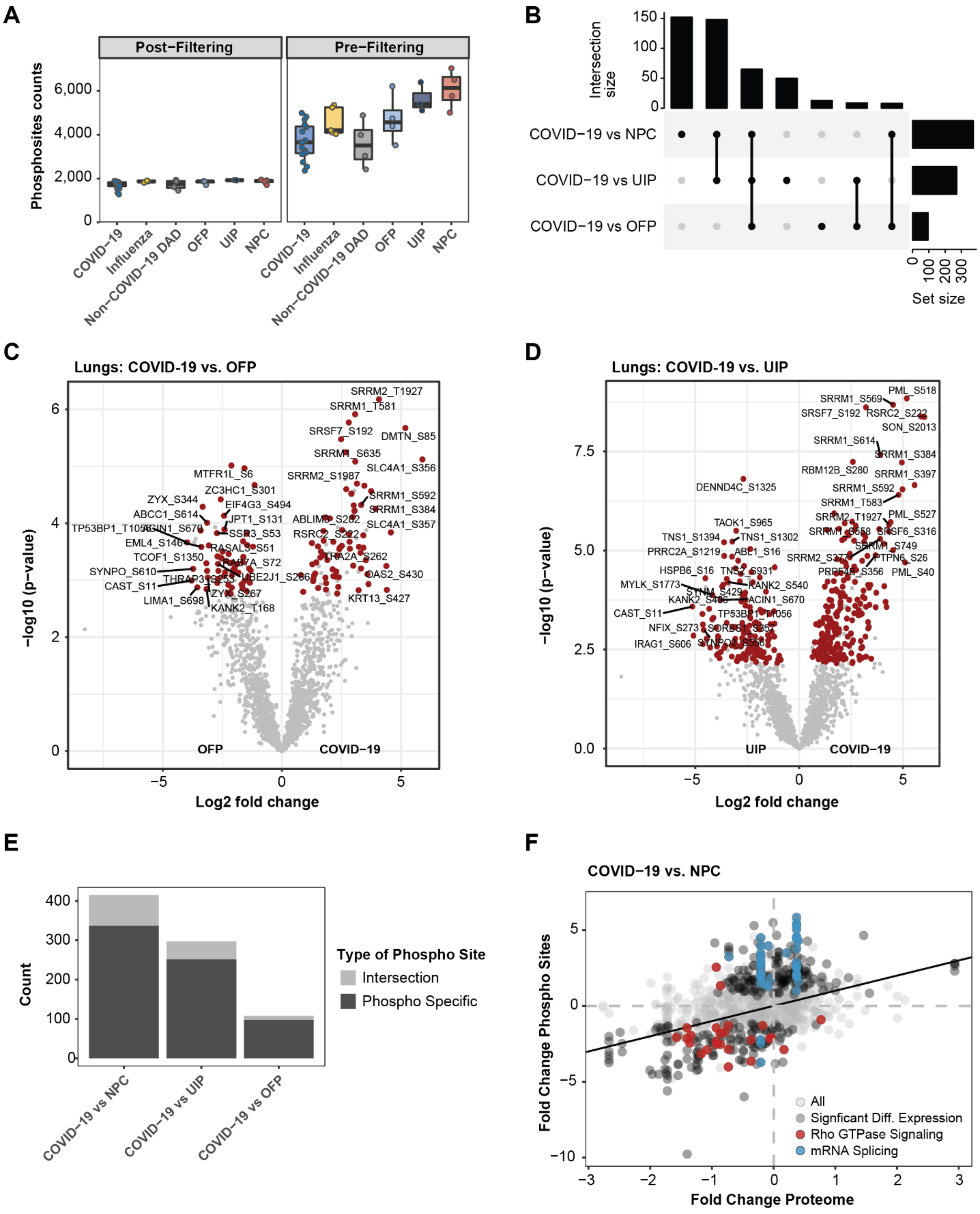
A
Post−Filtering
Pre−Filtering
Phosphosites counts
6,000
4,000
2,000
0
COVID−19
Influenza
Non−COVID−19 DAD
OFP
UIP
NPC
B
Intersection size
150
100
50
0
COVID−19 vs NPC
COVID−19 vs UIP
COVID−19 vs OFP
0
100
200
300
Set size
C
Lungs: COVID-19 vs. OFP
−log10 (p−value)
Log2 fold change
OFP
COVID−19
SRRM2_T1927
SRRM1_T581
SRSF7_S192
DMTN_S85
SRRM1_S635
SLC4A1_S356
MTFR1L_S6
SRRM2_S1987
ZC3HC1_S301
ZYX_S344
EIF4G3_S494
SRRM1_S592
SRRM1_S384
ABCC1_S614
JPT1_S131
ABLIM3_S262
SLC4A1_S357
SSR3_S53
RSRC2_S222
EML4_S146
RASAL3_S51
TCOF1_S1350
TRA2A_S262
SYNPO_S610
UBE2J1_S266
OAS2_S430
CAST_S11
ZYX_S267
KRT13_S427
LIMA1_S698
KANK2_T168
D
Lungs: COVID-19 vs. UIP
−log10 (p−value)
Log2 fold change
UIP
COVID−19
7.5
5.0
2.5
0.0
PML_S518
SRRM1_S569
SRSF7_S192
RSRC2_S222
SON_S2013
SRRM1_S614
SRRM1_S384
RBM12B_S280
SRRM1_S397
DENND4C_S1325
SRRM1_S592
SRRM1_T583
TAOK1_S965
SRRM2_T1927
PML_S527
TNS1_S1394
TNS1_S1302
SRSF6_S316
PRRC2A_S1219
ABL1_S16
SRRM1_S749
SRRM2_S377
PTPN6_S26
HSPB6_S16
PML_S40
MYLK_S1773
KANK2_S540
ACIN1_S670
CAST_S11
NFIX_S273
IRAG1_S606
E
Count
400
300
200
100
0
Type of Phospho Site
Intersection
Phospho Specific
COVID−19 vs NPC
COVID−19 vs UIP
COVID−19 vs OFP
F
COVID−19 vs. NPC
Fold Change Phospho Sites
Fold Change Proteome
5
0
−5
−10
−3
−2
−1
0
1
2
3
All
Signficant Diff. Expression
Rho GTPase Signaling
mRNA Splicing

#### Supplementary Figure 4

**A** Counts of quantified phosphorylation sites in COVID-19 and all control groups. Data are shown before (right) and after (left) stringent filtering as described (Material and Methods).

**B** Upset plot depicting the intersection of differentially expressed proteins across between COVID-19 and NPC, OFP or UIP, respectively. Statistical testing in comparison to Influenza and non-COVID-19 DAD did not result in significant hits.

**C, D** Differential phospho-site regulation between COVID-19 and OFP (**C**) or UIP (**D**), respectively. Protein significance (t-test, q-val < 0.05, fold change >1.5) is highlighted in red.

**E** Count of phospho-sites separated into unique differential alteration on the level of protein phosphorylation (Phospho Specific) compared to the portion of sites which correspond to significant differential expression on the protein level (Intersection).

**F** Comparison of fold changes between COVID-19 and NPC on the phosphorylation and whole proteome level. Proteins associated to Rho GTPase signaling (red) and mRNA splicing (blue) are highlighted; significant differential expression (t-test, q-val < 0.05, fold change >1.5) is indicated in grey.

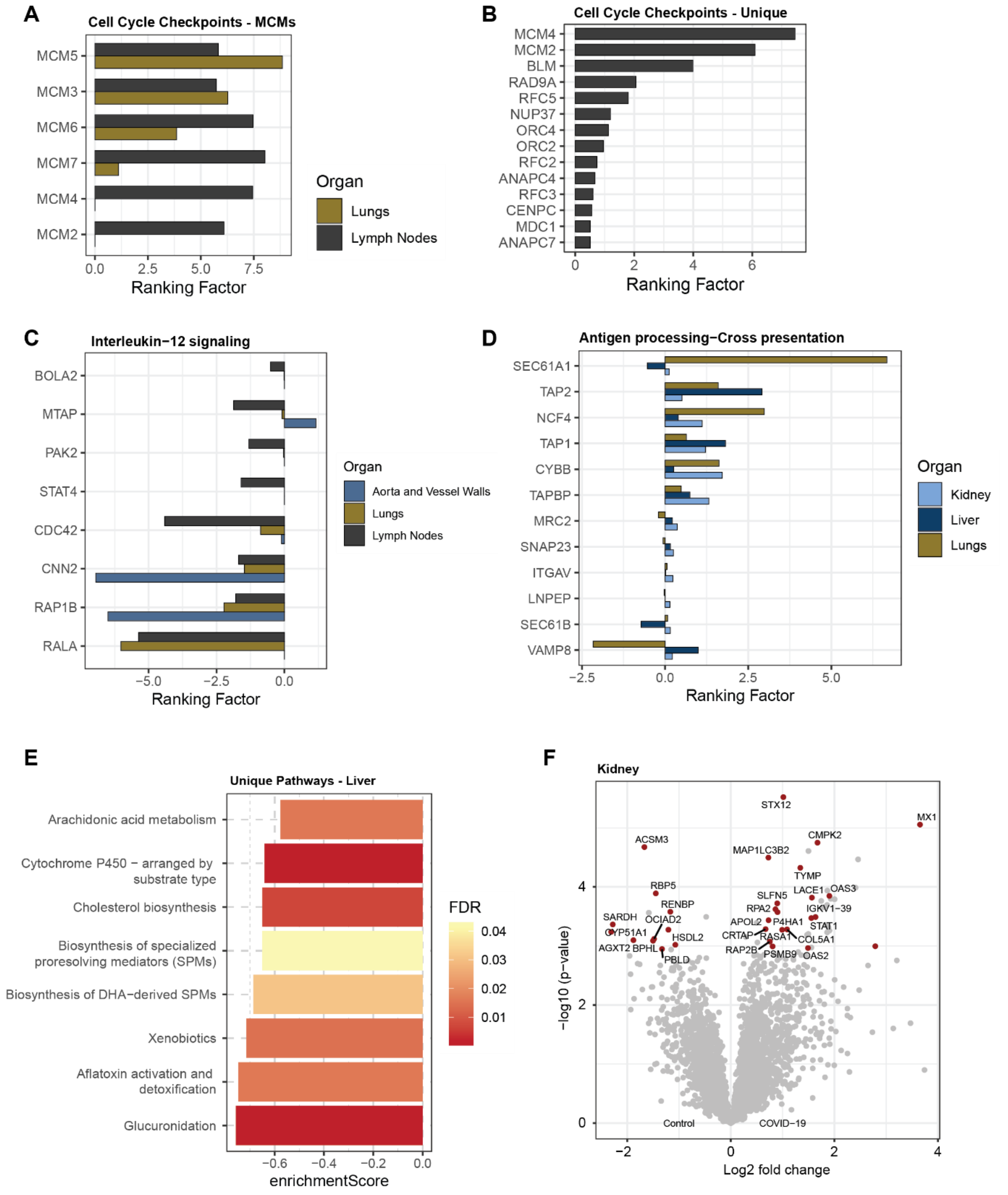
A
Cell Cycle Checkpoints - MCMs
MCM5
MCM3
MCM6
MCM7
MCM4
MCM2
0.0
2.5
5.0
7.5
Ranking Factor
Organ
Lungs
Lymph Nodes
B
Cell Cycle Checkpoints - Unique
MCM4
MCM2
BLM
RAD9A
RFC5
NUP37
ORC4
ORC2
RFC2
ANAPC4
RFC3
CENPC
MDC1
ANAPC7
0
2
4
6
Ranking Factor
C
Interleukin−12 signaling
BOLA2
MTAP
PAK2
STAT4
CDC42
CNN2
RAP1B
RALA
−5.0
−2.5
0.0
Ranking Factor
Organ
Aorta and Vessel Walls
Lungs
Lymph Nodes
D
Antigen processing−Cross presentation
SEC61A1
TAP2
NCF4
TAP1
CYBB
TAPBP
MRC2
SNAP23
ITGAV
LNPEP
SEC61B
VAMP8
−2.5
0.0
2.5
5.0
Ranking Factor
Organ
Kidney
Liver
Lungs
E
Unique Pathways - Liver
Arachidonic acid metabolism
Cytochrome P450 − arranged by substrate type
Cholesterol biosynthesis
Biosynthesis of specialized proresolving mediators (SPMs)
Biosynthesis of DHA−derived SPMs
Xenobiotics
Aflatoxin activation and detoxification
Glucuronidation
−0.6
−0.4
−0.2
0.0
enrichmentScore
FDR
0.04
0.03
0.02
0.01
F
Kidney
STX12
MX1
ACSM3
CMPK2
MAP1LC3B2
TYMP
RBP5
LACE1
OAS3
SLFN5
RENBP
RPA2
IGKV1−39
SARDH
OCIAD2
APOL2
P4HA1
STAT1
CYP51A1
HSDL2
CRTAP
RASA1
COL5A1
AGXT2
BPHL
RAP2B
PSMB9
OAS2
PBLD
Control
COVID−19
−log10 (p−value)
0
2
4
−2
0
2
4
Log2 fold change

#### Supplementary Figure 5

**A** Bar plot representing the ranking factor values (q-value (-log10) x fold change (log2)) of all MCM proteins associated to the Pathway term 'Cell Cycle Checkpoints' (Reactome database) commonly identified in lungs and lymph nodes.

**B** Ranking factor for proteins associated to the Pathway term 'Cell Cycle Checkpoints' (Reactome database) that have been uniquely identified in the lymph nodes.

**C** Proteins of Interleukin-12 signaling (Reactome database) in lungs, lymph nodes and aorta/vessel walls depicted by ranking factors.

**D** Bar plot of ranking factors for the pathway antigen processing-cross presentation (Reactome database) for lungs, liver and kidney. For reason of clarity, proteasomal subunits are excluded.

**E** Unique pathway terms for the liver which were identified using a GSEA enrichment (Reactome database). Color code of each bar indicate the FDR of the enrichment.

**F** Volcano plots of differential protein expression between COVID-19 control specimen in the kidney. Protein significance (t-test, q-val $< 0.05$, fold change $>1.5$) is highlighted in red.

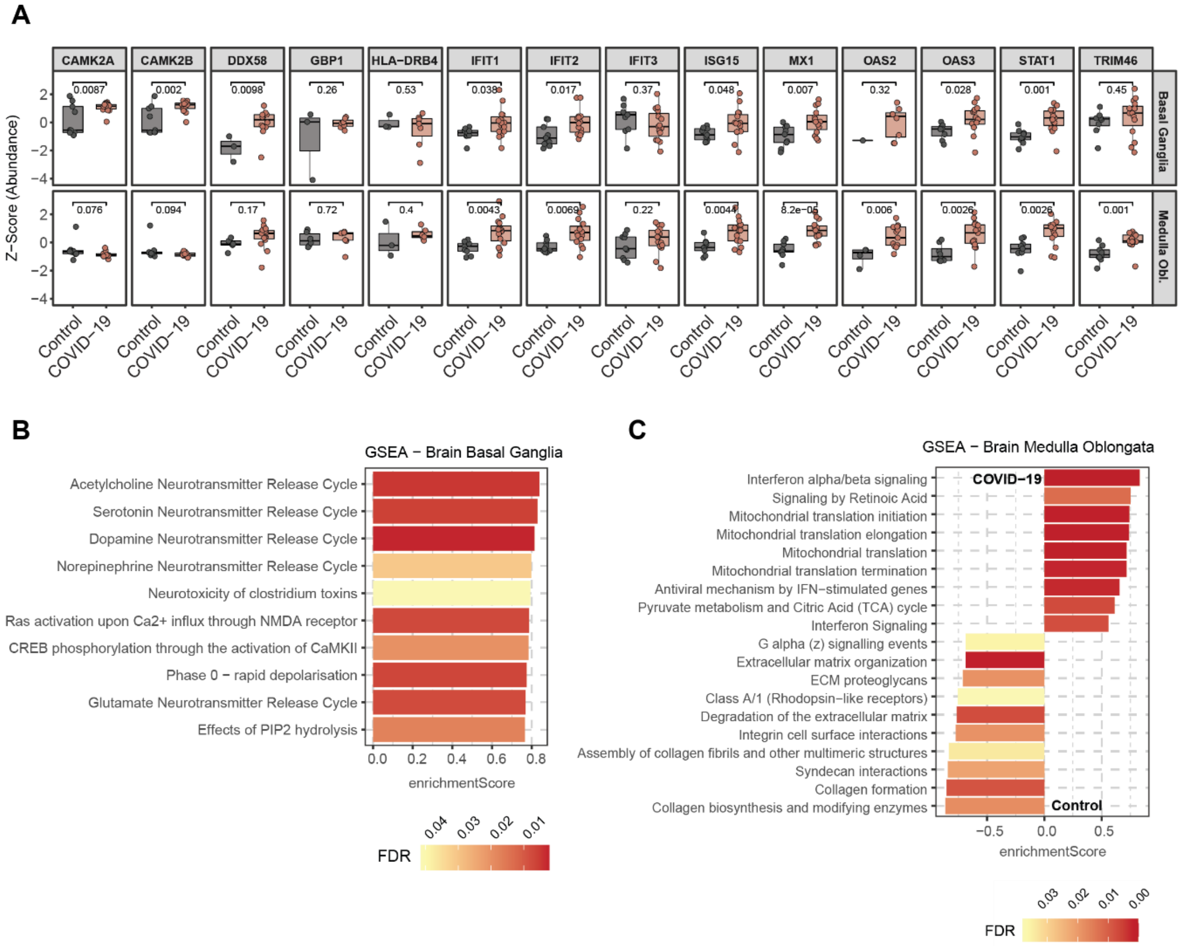


#### Supplementary Figure 6

**A** Z-Score protein abundance of proteins associated to 'Interferon Signaling' (Reactome database) that showed overlapping occurrence in both regions of the brain. An assessment of statistical significance (unpaired t-test) between COVID-19 and control samples is annotated for each comparison.

**B,C** GSEA enrichment depicted by the top 10 most significant terms assigned by enrichment score. A positive enrichment score indicates positive protein abundance in COVID-19, whereas a negative value indicates an enrichment in the control samples. The color coding of all bars depicts the FDR value of the respective enrichment term.

#### References

Coscia, F., Doll, S., Bech, J.M., Schweizer, L., Mund, A., Lengyel, E., Lindebjerg, J., Madsen, G.I., Moreira, J.M., and Mann, M. (2020). A streamlined mass spectrometry-based proteomics workflow for large-scale FFPE tissue analysis. J Pathol *251*, 100-112.
